# *SLC27A2* as a molecular marker of impaired epithelium in chronic rhinosinusitis with nasal polyps

**DOI:** 10.1101/2024.08.07.24311531

**Authors:** Jaewoo Park, Jung Yeon Jang, Jeong Heon Kim, Se Eun Yi, Yeong Ju Lee, Myeong Sang Yu, Yoo-Sam Chung, Yong Ju Jang, Ji Heui Kim, Kyuho Kang

## Abstract

**Background:** Chronic rhinosinusitis with nasal polyps (CRSwNP) is a complex disease characterized by multiple inflammatory endotypes. Although recent progress has been made in endotype-based classification, developing tailored therapeutic strategies for CRSwNP remains challenging. This study aimed to optimize therapeutic outcomes in CRSwNP by identifying potential molecular markers.

**Methods:** We utilized an integrated approach that combined bulk and single-cell RNA sequencing (scRNA-seq) to delineate the molecular signatures inherent to the cellular components of nasal polyp (NP) tissue. The levels of C11-BODIPY (as a marker of lipid peroxidation) and *SLC27A2*/FATP2 were assessed using quantitative PCR and immunofluorescence (IF) staining. The effects of lipofermata, a FATP2 inhibitor, were examined in air-liquid interface (ALI) cultured epithelial cells derived from CRSwNP patients and healthy controls.

**Results:** Deconvolution analysis of NP tissue revealed an upregulation of genes associated with lipid metabolism in the NP epithelium. In CRSwNP patients, we observed a significant increase in lipid peroxidation and *SLC27A2*/FATP2 expression in the NP epithelium. A marked expression of genes critical to metabolic pathways involved in lipid peroxidation was identified in *SLC27A2*-positive epithelial cells. Additionally, FATP2 and lipid peroxidation staining patterns exhibited a positive correlation in their respective % Area levels. Elevated *SLC27A2* expression was associated with disease pathogenesis and correlated with disease severity. Treatment with lipofermata resulted in decreased mRNA levels of *ALOX15*, a key mediator of inflammation and lipid peroxidation, and *FOXJ1*, a marker of abnormal ciliogenesis.

**Conclusion:** Elevated *SLC27A2* expression in the NP epithelium correlates with the severity of CRSwNP, highlighting its potential as a therapeutic target for managing advanced CRSwNP cases.

## 1. INTRODUCTION

Chronic rhinosinusitis (CRS) is a prevalent disease affecting up to 11% and 12% of the population in Europe, South Korea, and the United States, respectively, significantly impairing quality of life by presenting symptoms such as nasal congestion, discharge, facial pressure, anosmia, cough, and fatigue^1–3^ CRS is traditionally classified into two distinct phenotypes: chronic rhinosinusitis with nasal polyps (CRSwNP) and chronic rhinosinusitis without nasal polyps (CRSsNP).^2^ The endotypes of CRSwNP are determined by the type of inflammation present, including eosinophilic CRSwNP (ECRSwNP; type 2), and non-eosinophilic CRSwNP (nECRSwNP; non-type 2).^3,4^ Type 2 CRSwNP is predominant in Western populations, accounting for 80% of cases, while in Asian countries like Korea, Japan, and China, types 1 and 3 are more prevalent, constituting 20–60% of CRSwNP cases.^4–7^ Recent evidence has elucidated the complex interplay between various inflammatory pathways and tissue remodeling factors, delineating their relevance to the endotypes of CRSwNP.^8^ This growing understanding underscores the importance of a personalized, endotype-targeted approach to the management of CRSwNP.^3,9^ Despite these advances, however, the selection of an optimal therapeutic strategy for CRSwNP remains fraught with challenges, particularly due to the paucity of molecular biomarkers that adequately reflect the clinical characteristics of the various CRSwNP types. Moreover, the existence of cases with mixed or indistinct endotype-specific features exacerbates the intricacy of treatment decision-making.^7,9–13^ Consequently, CRSwNP remains a significant clinical conundrum in the quest for effective, customized therapeutic interventions.

Recent advances in genomic profiling have shed light on the pathogenesis of CRSwNP, providing potential solutions to the challenges faced by affected individuals. Whole-transcriptome sequencing has revealed gene signatures linked to inflammation and abnormal host defense mechanisms, as well as distinct transcriptomic differences between ECRSwNP and nECRSwNP.^14–16^ Single-cell transcriptome profiling has further enriched our understanding by revealing the complex cellular landscape within NP tissue, identifying subpopulations of immune and epithelial cells that contribute to type 2 immunity through the expression of pro-inflammatory mediators.^14,17–22^ Imbalances in cellular populations, such as basal cell hyperplasia and the loss of ciliated/secretory cells, have also been documented.^17,21^

Recent focus has turned to the role of lipid and fatty acid metabolic dysregulation in the pathogenesis of CRSwNP.^19–22^ *ALOX15*, a lipid mediator implicated in inflammation, has been observed in select subpopulations of macrophages and dendritic cells. Additionally, discernible expression of lipid transport-related genes—*PLTP*, *APOE*, *ABCA1*, and *LRP1*—has been identified within specific subunits of macrophages.^21^ Dysregulated fatty acid metabolism has been observed in nasal polyp-derived eosinophils.^23^ Similarly, T helper 2 cells exhibit an enrichment of genes associated with lipid metabolism.^20^ Imbalances in lipid metabolism can also induce epithelial dysfunction. In tuft cells within the NP epithelium, genes related to the arachidonic acid metabolism pathway, including *PTGS1* and *ALOX5*, have been found to be uniquely enriched.^19^ Despite its pro-inflammatory effects, *ALOX15*, a marker of lipid peroxidation, has been previously reported to exhibit robust expression in the NP epithelium.^24–31^ Recent findings have highlighted the presence of genes associated with lipid peroxidation in the NP epithelium.^32–34^ Despite these pivotal studies, our understanding of lipid metabolism in epithelial dysfunction remains incomplete, necessitating further research that encompasses all endotypes.

In this study, we aimed to identify molecular markers associated with aberrant epithelial activity that may serve as potential prognostic and therapeutic targets for patients with CRSwNP. We analyzed the cellular composition of NP tissues through deconvolution analysis of bulk and single-cell RNA-seq datasets. Our results demonstrated an increase in the expression of genes related to lipid metabolic processes in the epithelial cells of NPs. We found that fatty acid transporter protein 2 (FATP2), a long-chain fatty acid transporter encoded by the solute carrier family 27-member 2 (*SLC27A2*) gene, was significantly upregulated in NP epithelium compared to healthy control epithelium. Patients with high expression of *SLC27A2* exhibited increased mucociliary dysfunction-related transcriptional signature genes and severity scores. Inhibition of *SLC27A2* resulted in decreased expression of epithelial dysfunction-related genes. Our findings suggest that *SLC27A2* may be a novel prognostic and therapeutic target gene for patients with severe CRSwNP.

## 2. METHODS

### 2.1 Patient recruitment

We enrolled adult Korean participants with chronic rhinosinusitis with nasal polyps (CRSwNPs) and healthy controls from the Department of Otorhinolaryngology – Head and Neck Surgery at Asan Medical Center (AMC) between July 2019 and April 2022. Written informed consent was obtained from all subjects before the study’s commencement, conducted in accordance with the Declaration of Helsinki. This study received approval from the Asan Medical Center Institutional Review Board (IRB No. 2019-0619).

For patients with CRSwNPs, nasal polyp (NP) tissues were collected during functional endoscopic sinus surgery, and diagnoses followed the criteria outlined in the 2020 European Position Paper on Rhinosinusitis and Nasal Polyps guidelines, which included assessing symptoms, nasal endoscopy findings, and computed tomography (CT) imaging.^2^ Exclusion criteria included patients: 1) below 18 years; 2) with a history of unilateral CRS, antrochoanal polyps, fungal sinusitis, allergic fungal sinusitis, aspirin-exacerbated respiratory disease, cystic fibrosis, or primary ciliary dyskinesia; 3) who are pregnant or immunocompromised; 4) administered decongestants, antibiotics, and systemic/topical corticosteroids over four weeks preceding surgery; and 5) with a history of acute respiratory infection within four weeks prior to surgery.

Histopathological examination was performed to rule out fungal sinusitis, allergic fungal sinusitis, cystic fibrosis, or primary ciliary dyskinesia. We also inquired about their history of acute upper and lower respiratory tract reactions to aspirin or nonsteroidal anti-inflammatory drugs to identify aspirin-exacerbated respiratory disease. Additionally, we conducted various assessments, including the Sinonasal Outcome Test (SNOT-22), Lund-Mackay CT scores, Lund-Kennedy endoscopic score, nasal polyp score, and polyp grading according to the Davos classification.^2,35,36^

Control tissue samples were obtained from the middle turbinate (MT) mucosa of patients above 18 years old undergoing septoplasty for nasal obstruction without sinusitis on CT or transsphenoidal approach surgery for non-functioning pituitary tumors without adjacent structure invasion on MRI and sinusitis on CT scan. These control subjects had no history of asthma, upper respiratory infection, or administration of systemic, topical corticosteroids, or antibiotic medications within four weeks before surgery. We determined atopic status by detecting specific IgE antibodies to common inhalant allergens using ImmunoCAP™ tests (Phadia, Uppsala, Sweden), and asthma was diagnosed based on medical history and pulmonary function tests, including spirometry and challenge tests.

### 2.2 Utilization of publicly available data

In this study, we employed various publicly available datasets and databases as follows: Bulk RNA-seq datasets from Peng et al.^16^ and Nakayama, T., et al.^14^, single-cell RNA-seq datasets from Wang et al.^21^ and Ordovas-Montanes et al.^17^, and ATAC-seq data from Ordovas-Montanes et al.^17^

### 2.3 Bulk RNA-seq data analysis

For the analysis of bulk RNA-seq data (GSE136825; GSE179269), we mapped the reads to the hg38 genome using STAR version 2.7.1a with default parameters.^37^ Subsequently, we converted the aligned reads into tag directories using HOMER version 4.11.1.^38^ Quantification of each file was performed utilizing the “analyzeRepeats” script in HOMER. We normalized gene expression levels in each sample by calculating fragments per kilobase of transcript per million mapped reads (FPKM). Differential expression genes (DEGs) were identified through DESeq2 analysis based on raw read counts.^39^ The “getDifferentialExpression” command in HOMER was applied with criteria for significance defined as an adjusted p-value of < 0.05, more than a two-fold difference in expression levels, and an average FPKM > 2. Principal component analysis (PCA) distinctly classified the samples into Healthy and CRSwNP clusters. One Healthy sample, aligning more with the CRSwNP cohort, was recognized, and henceforth omitted from further assessments (GSE179269, Figure S2B).

### 2.4 Single-cell RNA-seq analysis

#### 2.4.1 Surgical biopsy of nasal polyp tissues

To profile the cellular ecosystem of nasal polyp (NP) tissues, we analyzed publicly available single-cell RNA-seq data (accession number HRA000772).^21^ To ensure compatibility with the publicly available bulk RNA-seq dataset (GSE136825) and to maintain uniformity in disease states and tissue locations, we utilized data from 11 samples obtained from surgical biopsies of nasal polyp tissues. These samples encompassed 5 non-ECRSwNP and 6 ECRSwNP patients.

For our analysis, we included only cells in which mitochondrial RNA accounted for less than 15% of the total RNA content. We performed data integration and anchoring using the top 10,000 highly variable features identified through the variance-stabilizing transformation method. Subsequently, we conducted uniform manifold approximation and projection (UMAP) analysis and constructed a nearest neighbor graph using the ‘FindNeighbors’ function. To identify cell clusters, we employed the ‘FindClusters’ function with a resolution parameter set to 0.5. Differential expression analysis was carried out using the ‘FindMarkers’ and ‘FindAllMarkers’ functions. Additionally, we assessed the expression levels of genes associated with lipid metabolism across various cell types using the ‘AddModuleScore’ function, which calculated module scores based on genes related to the GO:0006629 lipid metabolic process.

#### 2.4.2 Nasal scrapings

To identify cell populations expressing elevated levels of *SLC27A2*, we utilized previously published single-cell RNA-seq data (Ordovas-Montanes et al.).^17^ These data included 7,886 cells from the ethmoid sinus of 2 patients with chronic rhinosinusitis with nasal polyps (CRSwNP-NP), 2,031 cells from the inferior turbinate of 4 patients with CRSwNP (CRSwNP-IT), and 6,498 cells from the inferior turbinate of 3 healthy controls (Healthy-IT). We included only those cells in which mitochondrial RNA accounted for less than 20% of the total RNA content. Data integration and anchoring were performed using the top 3,500 highly variable features determined by the variance-stabilizing transformation method. Subsequently, we conducted uniform manifold approximation and projection (UMAP) analysis and constructed a nearest neighbor graph using the ‘FindNeighbors’ function. Cluster identification was achieved using the ‘FindClusters’ function with a resolution parameter set to 0.5. Differentially expressed genes (DEGs) were identified using the ‘FindMarkers’ and ‘FindAllMarkers’ functions. Principal component analysis and clustering were conducted using Seurat version 4.3.0.1 in R version 4.3.0.

### 2.5 Gene ontology analysis

To identify enriched Gene Ontology (GO) terms among the Differentially Expressed Genes (DEGs), we employed Metascape. ^40^

### 2.6 Module GSVA scores

For the assessment of variation in predefined gene sets across samples within our microarray expression datasets, we utilized Gene Set Variation Analysis (GSVA), specifically version 1.48.2, an open-source software available through R/Bioconductor.^41^ GSVA provides a non-parametric and unsupervised method for this purpose. The GSVA scores for each sample were calculated using the FPKM gene expression matrix of the selected gene set, and this computation was carried out using the GSVA package in R version 4.3.0.

### 2.7 Immunofluorescence

The samples (refer to Table 1) were fixed in 4% paraformaldehyde in phosphate-buffered saline (PBS) and subsequently embedded in paraffin using standard procedures. Sections of the specimens, each 4-μm in thickness, were subjected to heat-induced antigen retrieval by boiling in 10 mM citrate buffer (pH 6.0) and allowed to cool to room temperature for 20 minutes. The specimens were then incubated overnight at 4°C with anti-FATP2 (14048-1-AP, Proteintech, IL, USA; 1:100) antibodies. Afterward, the specimens were incubated with goat anti-rabbit Alexa Fluor 488 (A11008, Invitrogen, OR, USA; 1:200) and BODIPY™ 581/591 C11 (D3861, Invitrogen, OR, USA; 10 μM) for 2 hours at room temperature. Finally, the sections were counterstained with DAPI (D1306, Invitrogen, OR, USA; 1:1000) and examined using a confocal laser microscope (LSM 900, Carl Zeiss, Germany). Image analysis and quantification were performed using ImageJ version 1.53k.

**Table 1.** Demographics and clinical characteristics of subjects.

|  | Control | CRSwNP |
| --- | --- | --- |
| **RT-qPCR / comparison of severity score** | **20** | **20** |
| Age (y), mean ± SD | 38.1 ± 52.1 | 52.1 ± 12.9<sup>‡</sup> |
| Male, n (%) | 17 (85) | 18 (90) |
| Asthma, n (%) | 0 | 4 (20)<sup>§</sup> |
| Atopic, n (%) | 13 (65) | 7 (35) |
| SNOT-22, mean ± SD | 39.5 ± 17.5 | 42.1 ± 21.4 |
| Lund-Mackay CT score, mean ± SD | 0 | 18.8 ± 4.5 |
| Lund-Kennedy endoscopic score, mean ± SD | 0 | 9.7 ± 2.9 |
| Nasal polyp score, mean ± SD | 0 | 0.5 ± 2.2 |
| Polyp grade, mean ± SD | 0 | 3.7 ± 1.6 |
| **Immunofluorescence staining** | **3** | **16** |
| Age (y), mean ± SD | 54.3 ± 8.3 | 53.9 ± 12.5 |
| Male, n (%) | 2 (66.7) | 14 (87.5) |
| Asthma, n (%) | 0 | 3 (18.8) |
| Atopic, n (%) | 0 | 5 (31.3) |
| SNOT-22, mean ± SD | 8.7 ± 6.8 | 42.3 ± 21.1<sup>‡</sup> |
| Lund-Mackay CT score, mean ± SD | 0 | 19.3 ± 4.3<sup>‡</sup> |
| Lund-Kennedy endoscopic score, mean ± SD | 0 | 9.8 ± 2.8<sup>‡</sup> |
| Nasal polyp score, mean ± SD | 0 | 5.1 ± 2.4<sup>‡</sup> |
| Polyp grade, mean ± SD | 0 | 3.8 ± 1.7<sup>‡</sup> |
| **Oil-red O staining** | **3** | **7** |
| Age (y), mean ± SD | 61.7 ± 17.2 | 53.6 ± 20.8 |
| Male, n (%) | 1 (33.3) | 5 (71.4) |
| Asthma, n (%) | 0 | 2 (28.6) |
| Atopic, n (%) | 0 | 4 (57.1) |
| SNOT-22, mean ± SD | 3.7 ± 4.7 | 38.9 ± 21.9<sup>‡</sup> |
| Lund-Mackay CT score, mean ± SD | 0 | 16.9 ± 5.3<sup>‡</sup> |
| Lund-Kennedy endoscopic score, mean ± SD | 0 | 9.7 ± 2.4<sup>‡</sup> |
| Nasal polyp score, mean ± SD | 0 | 3.7 ± 1.8<sup>‡</sup> |
| Polyp grade, mean ± SD | 0 | 2.7 ± 0.9<sup>‡</sup> |
| **Lipofermata treatment** | **4** | **4** |
| Age (y), mean ± SD | 37.5 ± 10.3 | 44.25 ± 12.5 |
| Male, n (%) | 3 (75) | 4 (100) |
| Asthma, n (%) | 0 | 0 |
| Atopic, n (%) | 2 (50) | 0 |
| SNOT-22, mean ± SD | 14.8 ± 17.0 | 47.3 ± 34.4 |
| Lund-Mackay CT score, mean ± SD | 0 | 15.5 ± 4.8<sup>‡</sup> |
| Lund-Kennedy endoscopic score, mean ± SD | 0 | 8.8 ± 4.8<sup>‡</sup> |
| Nasal polyp score, mean ± SD | 0 | 8.8 ± 3.2<sup>‡</sup> |
| Polyp grade, mean ± SD | 0 | 3.5 ± 1.9<sup>‡</sup> |

‡ P < 0.05, Mann-Whitney U test
§, P < 0.05, chi-square test
SNOT, Sino-nasal outcome test

**Table 2.**
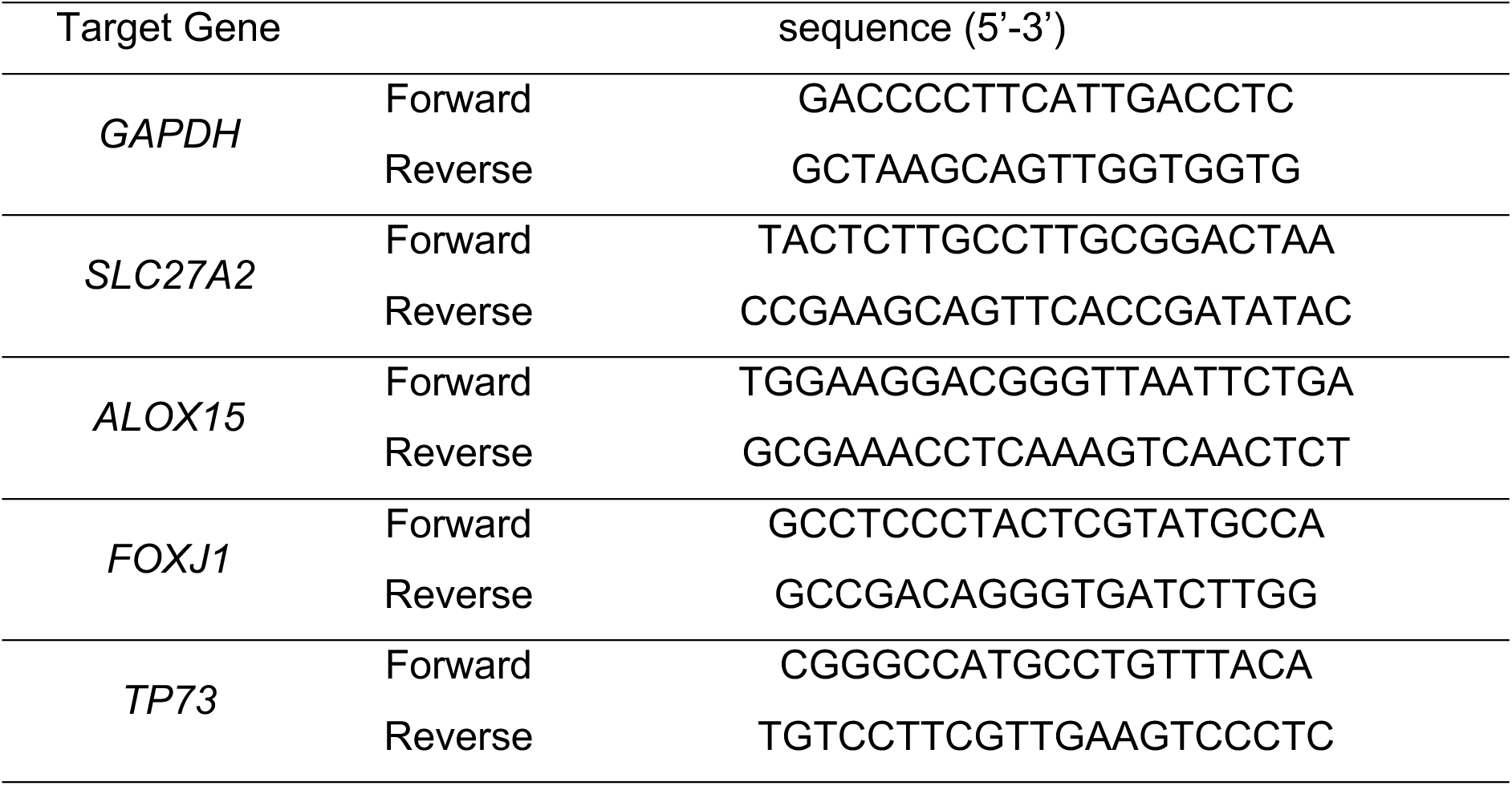
Primers for mRNA expression.

### 2.8 Oil Red O (ORO) staining

To assess lipid droplets in the tissue, we employed Oil Red O (ORO) staining. Fixed frozen tissue samples (see Table 1) were rinsed with 60% isopropanol for 2 minutes, followed by staining with a freshly prepared ORO working solution (Sigma-Aldrich, Catalogue #O1391) for 15 minutes. After staining, the sections were rinsed three times with 60% isopropanol (1 minute each) and twice with distilled water. Subsequently, sections were counterstained with 30% modified Mayer’s hematoxylin for 30 seconds, rinsed with tap water and two changes of distilled water, and finally, mounted in aqueous mounting media (Aquatex®, Sigma-Aldrich).

### 2.9 Quantitative real-time PCR

Total RNA was extracted from whole-tissue and epithelial cells of CRSwNPs and healthy controls (see Table 1) using the RNeasy Mini kit (QIAGEN). Reverse transcription was carried out using 500 ng of total RNA with the RevertAid First Strand cDNA Synthesis kit (Thermo Fisher Scientific). Real-time quantitative PCR (qPCR) was performed with TOPreal qPCR 2X PreMIX (SYBR Green with low ROX; No. RT500M; Enzynomics Co. Ltd., Daejeon, Korea). The qPCR conditions included an initial cycle at 95°C for 10 minutes, followed by 50 cycles of 10 seconds at 95°C for denaturation, 15 seconds at 60°C for annealing, and 20 seconds at 72°C for extension. A melting program was executed at 72–95°C with a heating rate of 1°C/45 seconds. The Rotor-Gene Q v. 2.3.1 (Qiagen, Hilden, Germany) was utilized to capture and analyze spectral data.

### 2.10 ATAC-seq analysis

For ATAC-seq analysis, sequenced reads were aligned to the human genome reference (GRCh38/hg38 assembly) using Bowtie2 version 2.3.4.1 with default parameters, and clonal reads were excluded from further analysis.^42^ A minimum of 10 million uniquely mapped reads was obtained for each condition. To identify peaks of ATAC-seq enrichment over the background, we utilized the ‘makeTagDirectory’ followed by the ‘findPeaks’ command from HOMER version 4.11.1.^38^ A false discovery rate (FDR) threshold of 0.001 was applied to all datasets. The total number of mapped reads in each sample was normalized to 10 million mapped reads. To visualize the normalized tag density of genes, we used the Integrative Genomics Viewer.

### 2.11 Air-liquid interface (ALI) culture

Primary human nasal epithelial cells (HNECs) were isolated from either polyp or control tissues (see Table 1). Passage-2 HNECs (1.5 × 105 cells/well) were seeded in 0.25 mL of culture medium on Transwell clear culture inserts (12 mm, with a 0.4 μm pore size; Costar; Corning Inc., Corning, NY). Initially, cells were cultured in a 1:1 mixture of basal epithelial growth medium and Dulbecco’s modified Eagle’s medium containing previously described supplements in a submerged state for the first 7–9 days. Upon reaching confluence, the apical medium was removed to establish an ALI, and thereafter, the medium was replenished only in the basal compartment. RNA was extracted from HNECs on day 14 following ALI establishment.^43,44^

### 2.12 In vitro lipofermata treatment

At day 14 of ALI culture, HNECs were exposed to either dimethyl sulfoxide (DMSO; SIGMA, 276855-100ML) or 2 μM lipofermata (MedChemExpress, HY-116788), a *SLC27A2*/FATP2 inhibitor, in the apical compartment for 24 hours. RNA was subsequently extracted from HNECs.

### 2.13 Statistical analysis

Unless otherwise specified, statistical analyses were performed using GraphPad Prism version 9 (GraphPad Software, La Jolla, CA, USA). Data are presented as mean ± standard error (SEM), unless otherwise stated in each figure legend. When applicable, two-tailed unpaired t-tests, paired t-tests, ordinary one-way ANOVA, and Wilcoxon signed-rank tests were employed. In all statistical analyses, similar expected variances were assumed between compared groups, and significance was accepted at the 95% confidence level (*P < 0.05, **P < 0.01, ***P < 0.001 for two-tailed unpaired t-tests, paired t-tests, and ordinary one-way ANOVA; *P < 0.05, **P < 0.01, ***P < 0.001 for Wilcoxon signed-rank tests).

## 3. RESULTS

### 3.1 Dissecting the nasal polyp transcriptome reveals altered lipid metabolism

To unveil unique molecular signatures within distinct cell types of NP tissues from CRSwNP patients, we employed a comprehensive transcriptomic approach, utilizing both bulk RNA-seq and scRNA-seq datasets. To enhance statistical robustness, we incorporated a publicly available bulk RNA-seq dataset (GSE136825; 28 Healthy, 42 CRSwNP).^16^ Additionally, we adopted a quality-assured public scRNA-seq dataset (HRA000772; 11 CRSwNP)^21^ to capture cell-type-specific molecular nuances (Figure 1A). Comparative analysis of the GSE136825 dataset revealed 971 differentially expressed genes (DEGs) between the 28 healthy and 42 CRSwNP samples. Among these DEGs, 552 were upregulated, and 419 were downregulated in CRSwNP (Figure S1A). Subsequent GO analysis shed light on potential molecular pathways and biological implications of these DEGs. Upregulated genes in CRSwNP were associated with GO terms such as ‘inflammatory response,’ ‘innate immune response,’ ‘cellular response to lipid,’ and ‘superoxide metabolic process,’ while downregulated genes were linked to ‘circulatory system process,’ ‘response to hormone,’ and ‘salivary secretion’ (Figure S1B). Using uniform manifold approximation and projection (UMAP), we identified eight distinct cell clusters encompassing 72,032 cells in the HRA000772 dataset. These clusters comprised epithelial cells, NK/T cells, B cells, myeloid cells, fibroblasts, endothelial cells, vascular smooth muscle cells (VSMCs), and cycling cells (Figure S1C). Cluster-specific marker genes were employed to delineate these cell types (Figure S1D).

**Figure 1.**
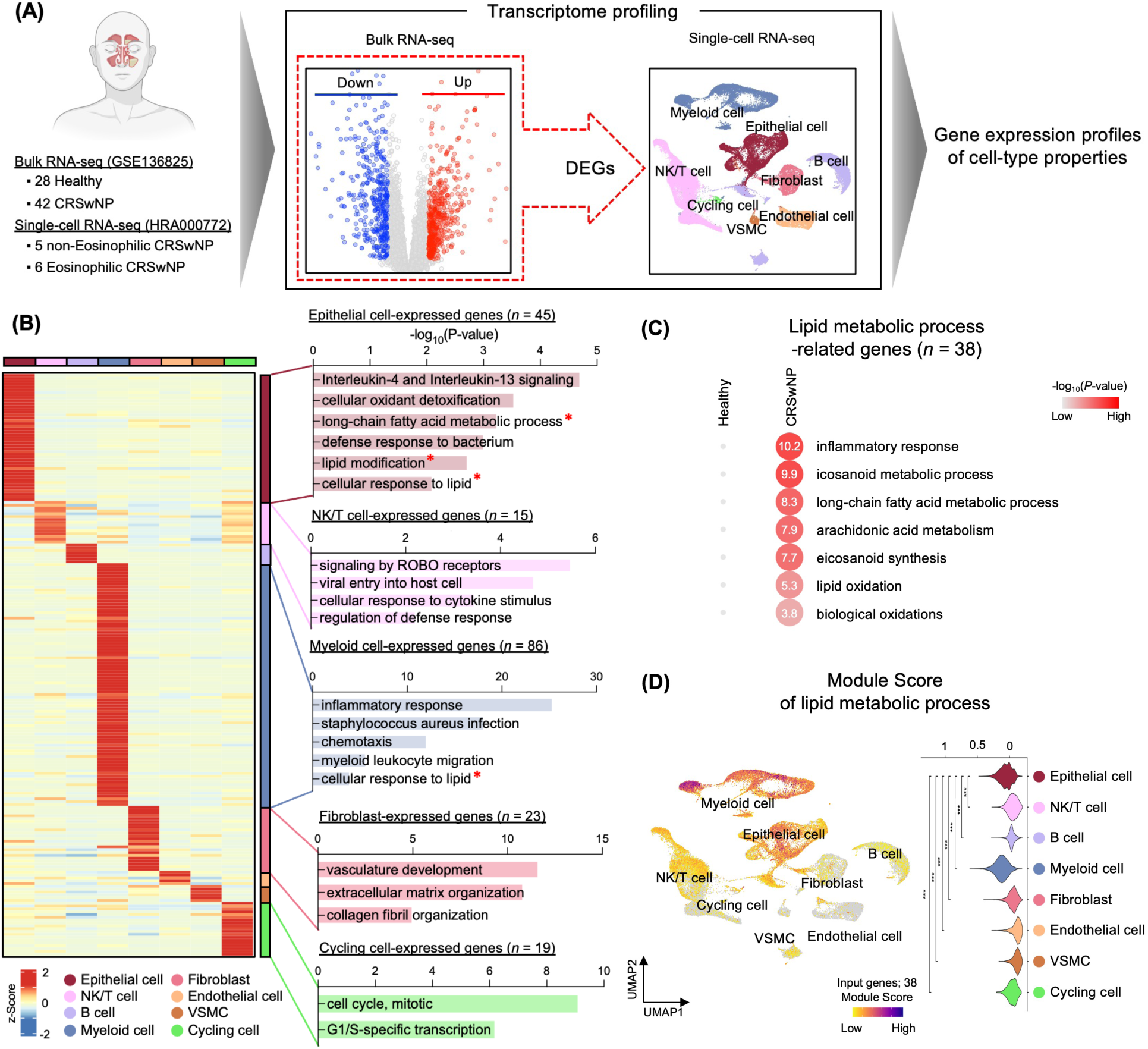
Transcriptomic profiling of the nasal polyp cellular microenvironment. (A) Schematic representation of the transcriptome profiling approach incorporating dataset GSE136825, comparing IT tissues from healthy controls (n = 28) to NP tissues from CRSwNP patients (n = 42). Dataset HRA000772 differentiates NP tissues of nECRSwNP patients (n = 5) from those with ECRSwNP (n = 6). Diagrams were produced using BioRender (https://biorender.com/). (B) Heatmap displaying the expression patterns of cell-type specific gene sets among upregulated DEGs in nasal polyp tissues, with values represented as z-scores (left). Gene ontology (GO) annotations for the marker gene sets associated with each distinct cell type are shown on the right. (C) Heatmap illustrating the enriched GO terms of lipid metabolic process-related genes upregulated in CRSwNP. (D) Module scores for lipid metabolic processes across individual cells are visualized on UMAP (left) and in a violin plot (right). Error bars represent mean values ± SEM. P-values in Figure (F) were determined using the Wilcoxon signed-rank test: * P < 0.05, ** P < 0.01, *** P < 0.001.

Subsequent integrated transcriptome analysis highlighted genes predominantly expressed in each cell type, with a particular focus on those upregulated in CRSwNP patients. These included 45 genes in epithelial cells, 15 in NK/T cells, 7 in B cells, 86 in myeloid cells, 23 in fibroblasts, 5 in endothelial cells, 6 in VSMCs, and 19 in cycling cells (Figure 1B, left). A subsequent GO analysis unveiled specific functional themes for these genes, elucidating characteristic GO terms for each cell type, such as ‘interleukin-4 and interleukin-13 signaling,’ ‘long-chain fatty acid metabolic process,’ and ‘lipid modification’ for epithelial cells, and ‘signaling by ROBO receptors’ for NK/T cells, among others (Figure 1B, right). Most of these GO terms aligned with established associations for each cell type in NP tissues. However, we gave priority to those GO terms with roles yet to be fully defined. Notably, lipid metabolism-linked GO terms featured prominently in the GO analysis of both whole tissues and specific cell types, especially epithelial and myeloid cells (Figure 1B). While previous studies have highlighted lipid metabolic processes in dendritic cells, macrophages, and Th2 cells^20,21^, their role in the NP epithelium remains an active area of investigation.

In our effort to comprehend the complex cellular milieu of nasal polyps, we first addressed potential analytical biases. Our primary objective was to identify genes associated with lipid metabolic pathways that exhibited elevated expression in CRSwNP patients. Through meticulous transcriptome analysis at the tissue level, we identified a set of 38 genes with increased expression in nasal polyp samples (refer to Figure S1E). Interestingly, pivotal genes such as *ALOX5, ALOX5AP, AOAH, APOE, PLA2G7*, and *TREM2* demonstrated marked upregulation in CRSwNP subjects compared to their healthy counterparts (Figure 1C). These genes were intricately linked to molecular processes and biological activities, particularly those related to inflammatory responses, arachidonic acid and eicosanoid metabolism, and lipid oxidation processes (Figure 1D). A deeper dive into the module scores of these 38 genes associated with lipid metabolic pathways revealed pronounced scores, primarily in myeloid, epithelial, and NK/T cells. Notably, epithelial cells exhibited an elevated module score, surpassed only by myeloid cells (Figures 1E and 1F). Our findings shed light on perturbed lipid metabolism in nasal polyp tissue, suggesting that these upregulated genes may be intricately involved in epithelial anomalies and heightened inflammatory events.

### 3.2 Augmented lipid peroxidation in nasal polyp epithelium of CRSwNP patients

To explore the potential association between lipid metabolic perturbations and atypical NP epithelium in CRSwNP patients, we correlated genes associated with lipid metabolic processes (38 genes) with those expressed in epithelial cells (45 genes). This alignment yielded 6 pertinent genes (Figure 2A). There was a marked difference in the mRNA expression levels of *ALOX15, GSTP1, IMPA2, SAA1, SGPP2*, and *SLC27A2* between the healthy and CRSwNP groups (Figure 2B). Furthermore, these genes exhibited robust expression in epithelial cell (Figure 2C and S2A). To reinforce our findings’ statistical robustness, we incorporated an external public bulk RNA-seq dataset (GSE179269; 7 Healthy, 17 CRSwNP; Figure S2B).^14^ In our exploration, CRSwNP patients had 1,290 upregulated genes and 580 downregulated genes. Significantly, of our identified 6 genes, *ALOX15, IMPA2, SGPP2*, and *SLC27A2* were conspicuously elevated in CRSwNP (Figure S2C and S2D). This skewed gene expression hints at disturbances in lipid metabolic processes, possibly pointing to lipid peroxidation and accumulation anomalies.^26,31,45–47^

**Figure 2.**
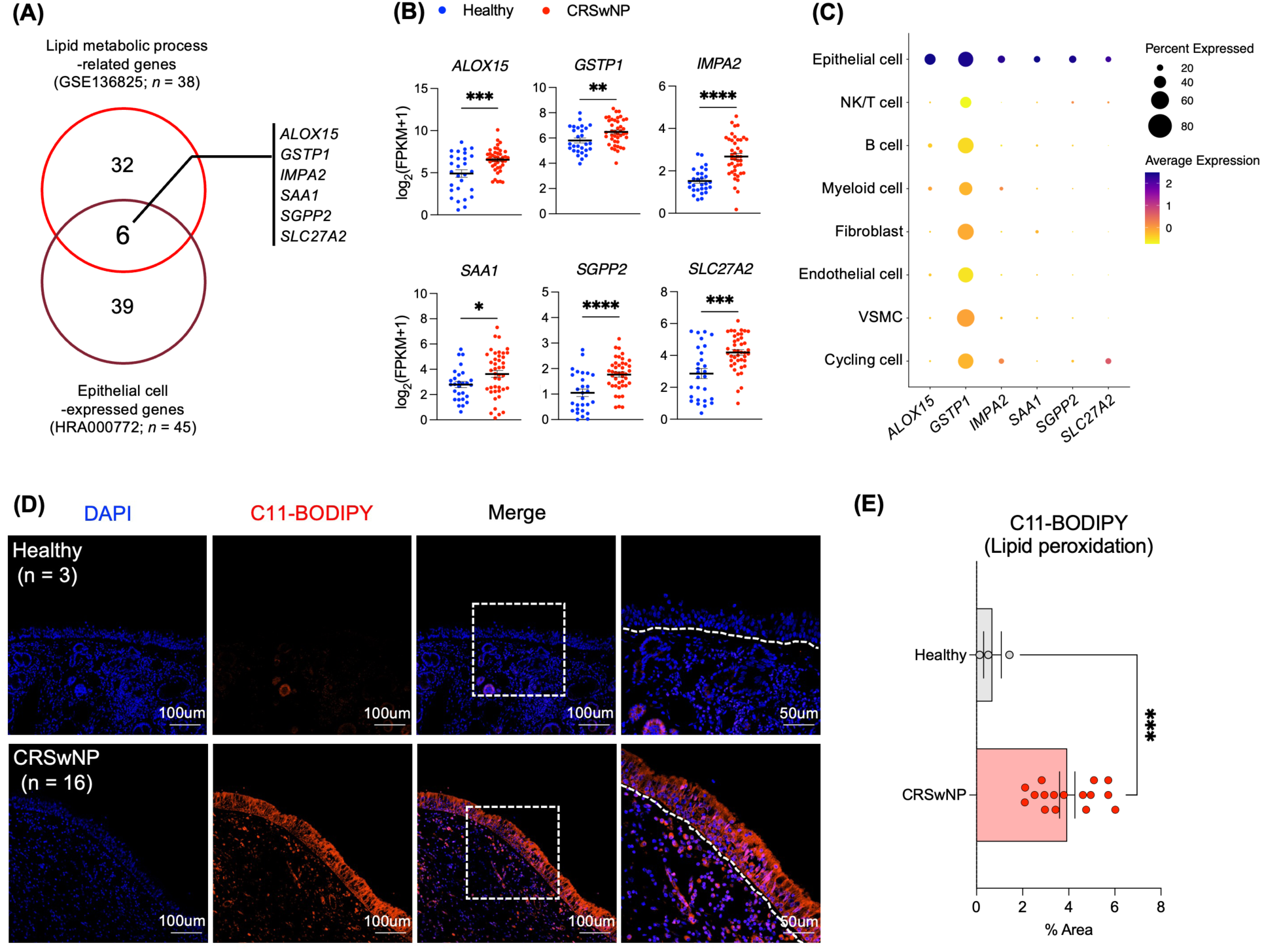
Enhanced lipid peroxidation in the nasal polyp epithelium of CRSwNP patients (A) Venn diagram depicting the overlap between upregulated lipid metabolic process-related genes in CRSwNP and those expressed in the epithelial cells of NP. (B) Expression levels of selected genes in CRSwNP relative to healthy controls, based on data from GSE136825. (C) Dot plot illustrating the specific cell types expressing the candidate genes. Color intensity represents the ratio of expression level in each cell cluster relative to the highest expression level across all clusters. Dot size correlates with the percentage of cells in each cluster expressing the gene. (D) Representative immunofluorescence staining images of C11-BODIPY (red) co-localized with DAPI (blue) in both healthy (n = 3) and CRSwNP samples (n = 16). (E) Quantification of the percentage area stained by C11-BODIPY, analyzed using ImageJ. Error bars represent mean values ± SEM. P-values were derived from an unpaired t-test. * P < 0.05, ** P < 0.01, *** P < 0.001, **** P < 0.0001. Scale bars: 50μm and 100μm.

Extending our preliminary assessments, we examined lipid accumulation and lipid peroxidation in NP tissues of both healthy controls and CRSwNP patients using Oil Red O (ORO) staining and C11-BODIPY (Figure S2E and Figure 2D). As illustrated in Figure 1D, CRSwNP patients exhibited a pronounced increase in lipid peroxidation relative to the healthy group (Figure 2D and 2E). Our data underscores the likelihood of upregulated genes involved in lipid metabolic dysfunctions in the NP epithelium, which might amplify epithelial irregularities, partially instigated by intensified lipid peroxidation.

### 3.3 Enhanced *SLC27A2*/FATP2 expression in nasal polyp epithelium

Fatty acid transporters play a critical role in lipid metabolism regulation. Disruption in their expression can significantly affect lipid homeostasis, possibly leading to various cellular abnormalities and diseases.^48–51^ Among the identified candidate genes, *SLC27A2* encodes for FATP2, a fatty acid transporter. Beyond *SLC27A2*, we assessed mRNA levels of other related genes including the solute carrier 27A (SLC27A) gene family, fatty acid binding proteins (FABPs), and CD36, contrasting their expressions between healthy individuals and CRSwNP patients. Notably, of the 16 genes examined, only *SLC27A2* exhibited a pronounced elevation in expression in CRSwNP (Figure 2B, 3A, S3A, and S3B). RT-qPCR validation further confirmed the marked difference in *SLC27A2* mRNA expression levels (Figure 3B). We subsequently investigated whether the alterations in *SLC27A2* expression were evident in nasal scrapings as well as in whole tissues.^17^ Cells expressing *SLC27A2* were notably present in epithelial cell clusters derived from CRSwNP nasal polyps (Figure 3C and 3D). In alignment with this, FATP2 levels were notably elevated in the NP epithelium of CRSwNP patients when compared to healthy controls (Figure 3E and 3F).

**Figure 3.**
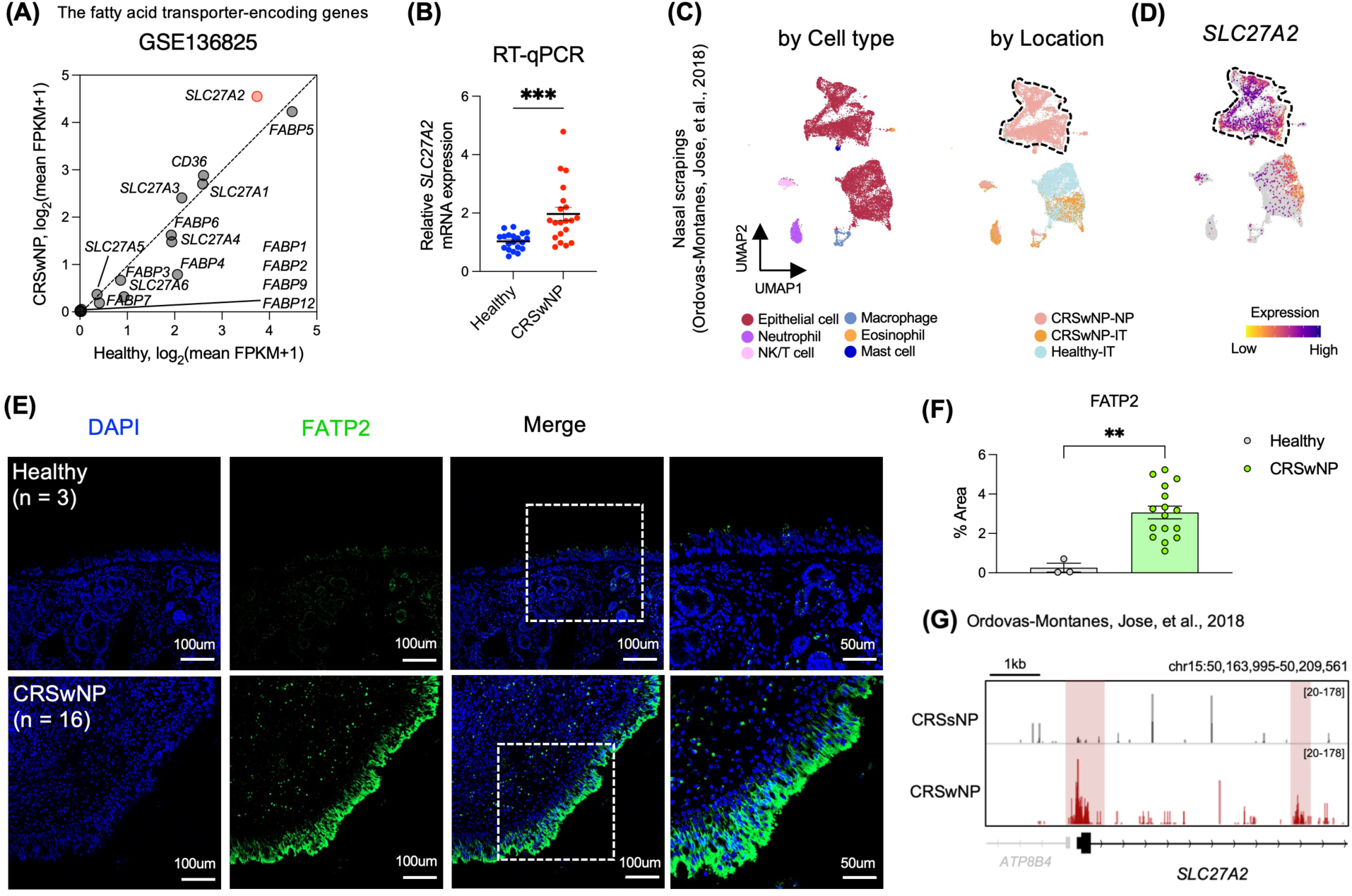
Enhanced expression of *SLC27A2*/FATP2 in nasal polyp epithelium (A) Scatter plot showcasing the disparities in gene expression levels of fatty acid transporter-encoding genes between the CRSwNP and healthy groups based on the GSE136825 dataset. (B) mRNA expression levels of *SLC27A2*, quantified via RT-qPCR and normalized to GAPDH mRNA, comparing CRSwNP (n = 20) and healthy samples (n = 20). (C) UMAP projection of previously published single-cell RNA-seq data (Ordovas-Montanes, Jose et al., 2018) comprising 16,415 individual cells, with color coding by cell identity. On the left is a UMAP representation of 16,415 cells derived from nasal scrapings (n = 9), categorized by cell type. The right displays a UMAP representation color-coded by disease location, covering samples directly scraped from the ethmoid sinus polyp in CRSwNP (7,886 cells, n = 2 CRSwNP-NP), directly scraped from the IT tissue of CRSwNP (2,031 cells, n = 4 CRSwNP-IT), and directly scraped from the IT tissue of a healthy control (6,498 cells; n = 3 Healthy-IT). (D) Gene expression distribution of *SLC27A2* visualized on feature plots. (E) Illustrative IF staining images of *SLC27A2*/FATP2 (shown in green) combined with DAPI (depicted in blue) for healthy (n = 3) and CRSwNP samples (n = 16). (F) Quantitative assessment of the area stained with anti-FATP2 using ImageJ software. (G) A representative IGV genome browser track, highlighting the normalized tag density associated with *SLC27A2*. Error bars indicate the mean value ± SEM. Significance was determined using an unpaired t-test: * P < 0.05, ** P < 0.01, *** P < 0.001, **** P < 0.0001. Scale bars are set at 50 μm and 100 μm.

Persistent chronic inflammatory stimuli can result in dynamic shifts in chromatin accessibility across all cellular types, inclusive of epithelial cells.^17,52–55^ Consequently, utilizing Omni-ATAC-seq data,^17^ we aimed to identify variations in the *cis*-regulatory components of genes implicated in lipid metabolic pathways, notably *SLC27A2*, within epithelial cells from nasal polyps. We observed increased chromatin accessibility in promoters and introns of genes linked to lipid metabolic processes (Figures 3G and S3C). Yet, However, these alterations exhibited no discernible differences among the endotypes of CRSwNP. A parallel analysis failed to discern notable differences in mRNA and protein expressions between ECRSwNP and nECRSwNP (Figures S3D, S3E, and S3F). Lipid peroxidation levels appeared consistent across both groups (Figures S3E and S3G). This observation was substantiated further by an external scRNA-seq dataset (HRA000772)^21^ incorporated in our research (Figures S3H, S3I, and S3J). Collectively, our findings indicate an elevated expression of *SLC27A2* in the epithelial cells of CRSwNP patients, irrespective of the endotype, possibly suggesting an epigenetic memory imprint.

### 3.4 Transcriptomic signatures and elevated lipid peroxidation in *SLC27A2*-positive nasal polyp epithelial cells

To elucidate the distinct molecular signature of *SLC27A2*-positive (*SLC27A2*+) cells in the NP epithelium, we employed the scRNA-seq dataset^21^ used for integrated transcriptome analysis (Figure 1). We isolated the epithelial cell cluster, subsequently dividing it into seven subtypes (Figure 4A). Each cluster-specific gene list identified the cell types of these subtypes (Figure S4A).

**Figure 4.**
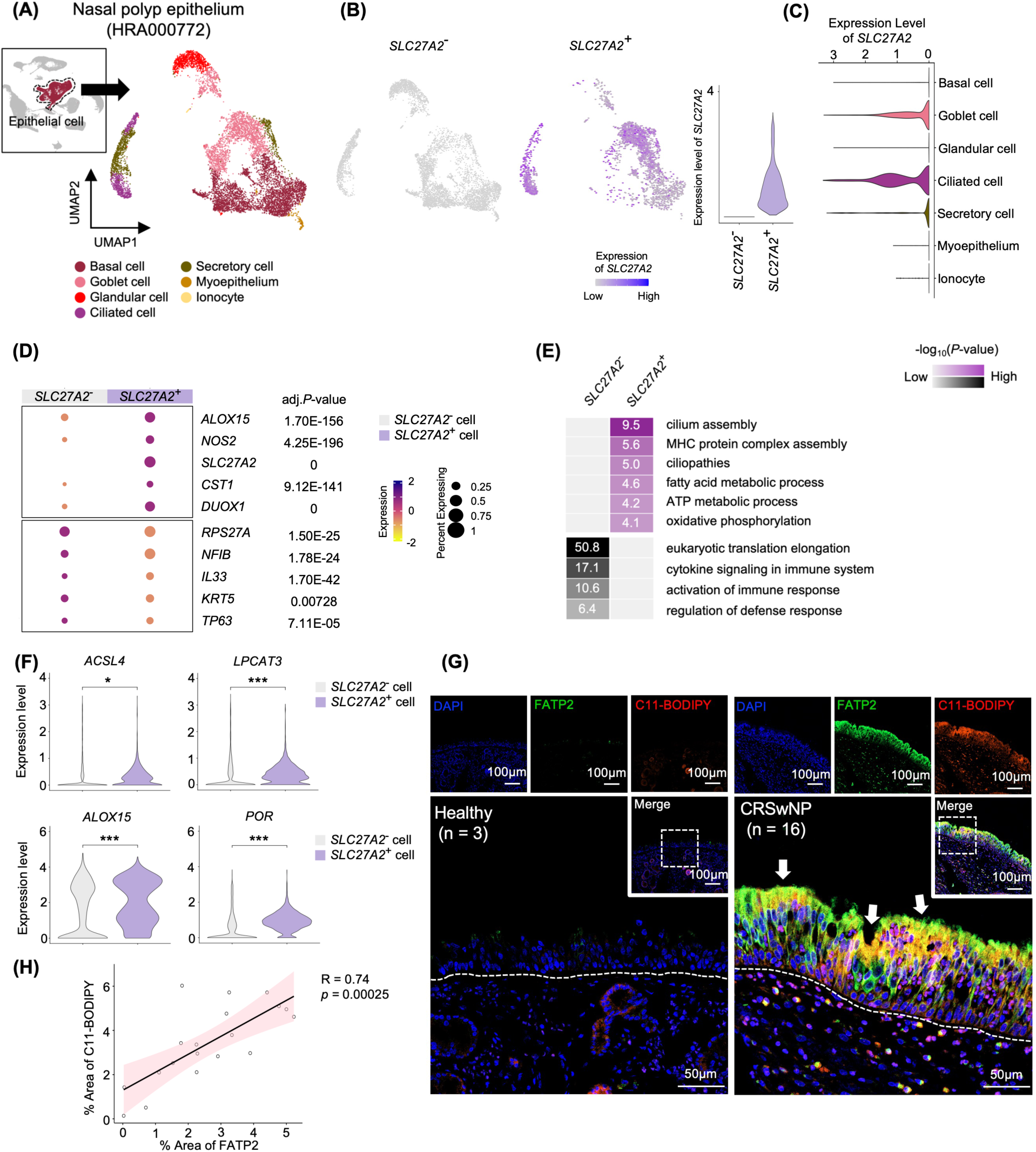
Unique transcriptomic signatures and enhanced lipid peroxidation in *SLC27A2*-positive nasal polyp epithelial cells (A) UMAP plot displaying 9,148 epithelial cells from 11 individuals (5 patients with nECRSwNP and 6 patients with ECRSwNP) categorized into 7 subsets. (B) UMAP plots depicting *SLC27A2*− and *SLC27A2*+ epithelial cells in nasal polyps from 11 patients with CRSwNP. *SLC27A2* expression is color-indicated (left). Violin plot showing the expression levels of *SLC27A2* in *SLC27A2*− and *SLC27A2*+ epithelial cells (right). (C) Violin plot illustrating the expression levels of *SLC27A2* across 7 epithelial subsets. (D) Dot plots showcasing representative gene expression levels in both *SLC27A2*− and *SLC27A2*+ epithelial cells. (E) Heatmap representing the enriched GO terms of differentially expressed genes (DEGs) between *SLC27A2*− and *SLC27A2*+ epithelial cells. Values are -log_10_(*P*-value). (F) Violin plot displaying the expression levels of *ACSL4, LPCAT3, ALOX15*, and *POR* in *SLC27A2*− and *SLC27A2*+ epithelial cells. (G) Representative immunofluorescence (IF) staining of *SLC27A2*/FATP2 (green) and C11-BODIPY (red) merged with DAPI (blue) in healthy tissue (n = 3) and CRSwNP tissue (n = 16). (H) Scatter plot depicting correlations between the percentage area stained by anti-FATP2 and C11-BODIPY. *P*-values and correlation coefficients (R) were determined using Spearman correlation analysis. Error bars represent mean values ± SEM. P-values were derived from the Wilcoxon signed-rank test. * *P* < 0.05, ** *P* < 0.01, *** *P* < 0.001. Scale bars: 50 μm and 100 μm.

Using a demarcation of 0 for *SLC27A2* expression, we classified epithelial cells into *SLC27A2*-negative (*SLC27A2*-) and those with an expression above 0 as *SLC27A2*+ (Figure 4B). Notably, the *SLC27A2*+ cells constituted 30.18% of the NP epithelium, with a predominance in the Goblet cell, Basal cell, Ciliated cell, and Secretory cell subtypes (Figure S4B). The distribution of *SLC27A2*+ cells showed a marked presence in Goblet cell (43.10%) and Ciliated cell (13.76%) compared to *SLC27A2*-cells (Goblet cell, 19.24%; Ciliated cell, 5.06%) (Figure S4C). Among the epithelial subtypes, the Ciliated cell exhibited the highest *SLC27A2* expression (Figure 4C).

Distinct gene expression patterns emerged between the two groups. Genes *ALOX15, NOS2, CST1,* and *DUOX1* consistently showed increased expression in *SLC27A2*+ cells, while *RPS27A, NFIB, IL33, KRT5,* and *TP63* were more expressed in *SLC27A2*-cells (Figure 4D). Gene Ontology (GO) analysis revealed unique transcriptomic features: the *SLC27A2*+ group showed increased genes linked to ciliopathies, MHC protein complex assembly, fatty acid metabolism, and oxidative phosphorylation, whereas the *SLC27A2*-group was enriched in genes related to cytokine signaling and immune response activation (Figure 4E).

In the cohort expressing *SLC27A2*, an elevated transcriptional response was observed, notably for genes involved in reactive oxygen species (ROS) metabolism and lipid oxidation upregulation (Figure S4D). Key enzymes like *ACSL4, LPCAT3, ALOX15*, and *POR*, pivotal for lipid peroxidation initiation, in conjunction with FATP2, showed marked expression disparities (Figure 4F).^26–29,31,56^ Immunofluorescence (IF) analysis revealed a concurrent localization between FATP2 and lipid peroxidation markers (Figure 4G). Additionally, a direct association was detected between FATP2 coverage area and lipid peroxidation extent (Figure 4H). This data underscores that, contrasting the heightened immune activity in *SLC27A2*-negative cells, the *SLC27A2*-positive subset, potentially linked to mucociliary clearance anomalies, manifests amplified lipid peroxidation events.

### 3.5 Differential transcriptomic characteristics of *SLC27A2*^High^ CRSwNP

To decipher the transcriptomic distinctions of CRSwNP patients with elevated *SLC27A2* expression, we analyzed the cohort from the GSE136825 study (n = 42, shown in Figure 1), applying a cutoff average value (log_2_FPKM = 4.18) to categorize subjects into ‘*SLC27A2*^Low^ CRSwNP’ and ‘*SLC27A2*^High^ CRSwNP’ groups (Figure 5A). A group of 21 healthy individuals provided a baseline for comparison. Profiling of gene expression delineated three distinct transcriptomic clusters, with 2,317 genes exhibiting differential expression among the groups. The ‘*SLC27A2*^High^ CRSwNP’ group (Cluster 1) manifested a significant differential expression of 844 genes. A further 661 genes were co-upregulated in both the ‘*SLC27A2*^Low^’ and ‘*SLC27A2*^High^’ CRSwNP clusters (Cluster 2), and 812 genes were observed to be downregulated in CRSwNP relative to the healthy control group (Cluster 3), as explicated in Figure 5B.

**Figure 5.**
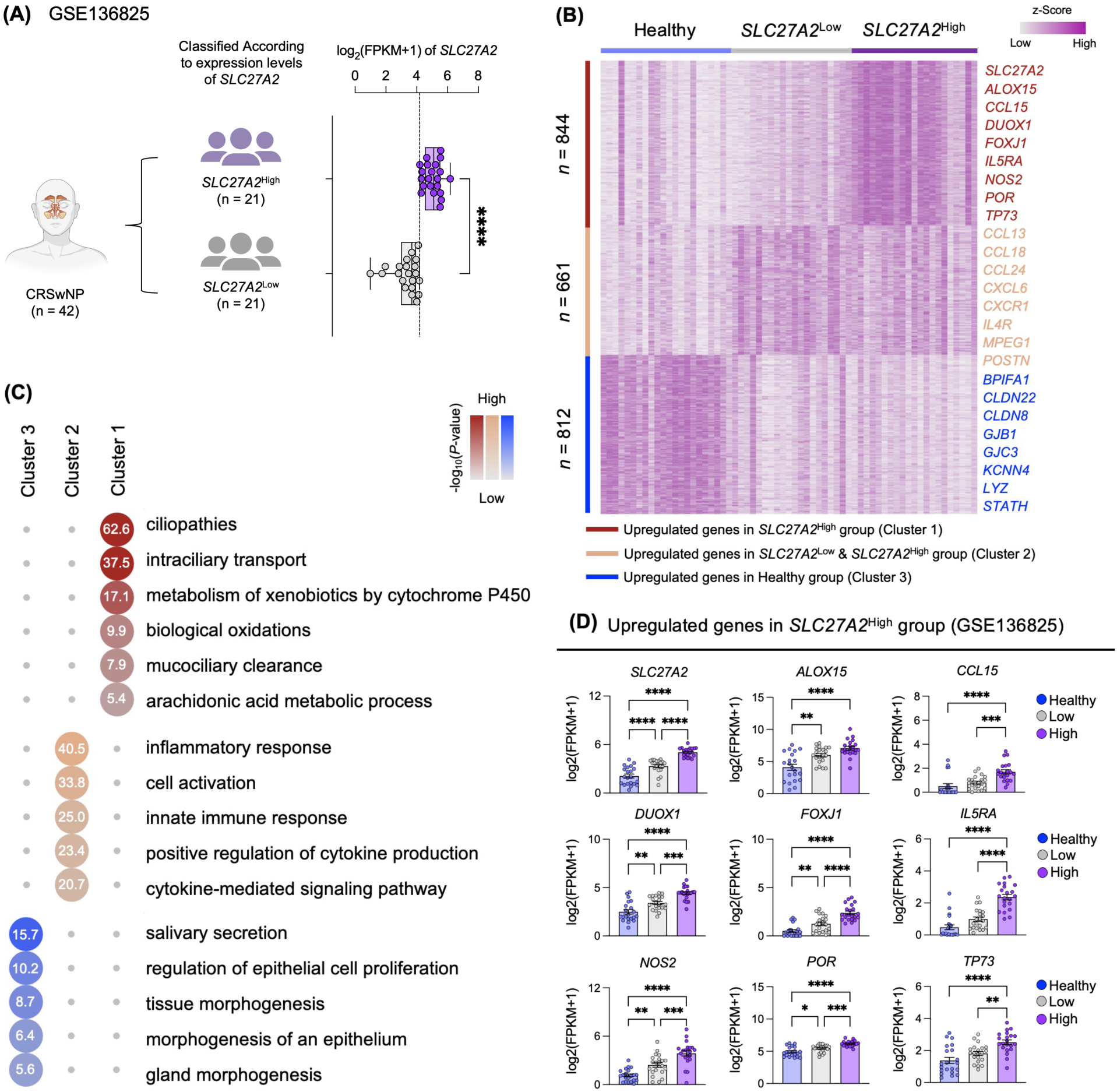
Transcriptomic features in CRSwNP with high *SLC27A2* expression (A) Schematic representation of the classification of CRSwNP patients based on *SLC27A2* expression levels using data from the GSE136825 dataset. Diagrams were created using BioRender (https://biorender.com/). (B) Heatmap depicting expression levels of differentially expressed genes (DEGs) (FDR < 0.05; two-fold difference in expression; average FPKM > 2). This analysis incorporates both hierarchical and k-means clustering (k = 3) of 2,317 DEGs among the three groups, based on data from GSE136825. (C) Heatmap of enriched Gene Ontology (GO) terms associated with each of the three clusters. Values are represented as –log_10_(*P*-value). (D) Comparative analysis highlighting the expression levels of select genes from Cluster 1, utilizing data from GSE136825. Error bars indicate mean values ± SEM. P-values were calculated using an unpaired t-test and ordinary one-way ANOVA. * P < 0.05, ** P < 0.01, *** P < 0.001, **** P < 0.0001.

Subsequent GO analysis provided insight into the functional characteristics of the identified clusters. Cluster 1 was characterized by genes involved in ciliopathies, biological oxidation, mucociliary clearance, and arachidonic acid metabolism. The GO terms of Cluster 2 underscored genes linked to the inflammatory response and immune system pathways, emblematic of CRSwNP’s inflammatory profile. In stark contrast, Cluster 3 was defined by genes associated with salivary secretion and the regulation of epithelial cell proliferation (Figure 5C). Genes corresponding to pivotal GO terms within each cluster were identified. To corroborate these insights, we cross-validated the gene signatures against an RNA-seq dataset from an independent cohort (GSE179269), detailed in Supplementary Figures 2 and 3.

Notably, Cluster 1, encompassing *SLC27A2*, included key genes such as *ALOX15*, which plays a role in inflammation and lipid peroxidation,^24,26–31^ as well as *DUOX1* and *POR*, essential for lipid peroxidation and the escalation of ROS.^57,58^ This cluster also contained *CCL15, IL5RA,* and *NOS2*, which are associated with airway inflammatory responses,^59–62^, alongside *FOXJ1* and *TP73,* identified as markers for atypical ciliary morphology and ciliogenesis (Figure 5D and S5A).^63^ Conversely, the signature genes of Cluster 2 included *CCL24, IL4R,* and *POSTN* (Figure S5B), whereas Cluster 3 was characterized by the expression of *GJC3, LYZ*, and *STATH* (Figure S5C). These findings articulate the distinct transcriptomic signatures of *SLC27A2*^High^ CRSwNP patients, emphasizing the profound roles of lipid peroxidation, ciliopathies, and inflammation in the pathology of the disease, offering a window into the molecular heterogeneity underlying CRSwNP.

### 3.6 Correlation between increased *SLC27A2* expression and CRSwNP severity

Investigating the correlation between *SLC27A2* expression and the severity of CRSwNP, we categorized patients from our cohort (outlined in Table 1) into two groups based on *SLC27A2* mRNA expression: ‘*SLC27A2*^Low^ CRSwNP’, encompassing 10 patients with lower expression levels, and ‘*SLC27A2*^High^ CRSwNP’, including 10 patients with higher expression. Clinical assessments utilizing the Lund-Mackay CT score, Lund-Kennedy endoscopic score, nasal polyp score, and polyp grade consistently demonstrated elevated indices in the ‘*SLC27A2*^High^ CRSwNP’ group (Figure 6A). Furthermore, a pronounced correlation emerged between elevated *SLC27A2* mRNA expression and increased levels of CRSwNP pathogenesis markers including *ALOX15*, *FOXJ1*, and *TP73*, primarily in the ‘*SLC27A2*^High^ CRSwNP’ group (Figure 6B and Figure S6A). Extending our investigation, an in vitro air-liquid interface (ALI) culture model of nasal epithelial cells was employed to discern the effect of *SLC27A2* inhibition on disease severity. Nasal epithelial cells derived from four healthy individuals and four CRSwNP patients were subjected to ALI culture. Following treatment with DMSO or 2 μM lipofermata, a *SLC27A2*/FATP2 inhibitor, we assessed the expression levels of pathogenesis-related markers *via* RT-qPCR (Figure 6C). Lipofermata treatment resulted in a significant reduction of *SLC27A2* expression in both normal and CRSwNP-derived epithelial cells (Figure 6D). Notably, *ALOX15* mRNA levels, associated with inflammatory processes, were significantly reduced in CRSwNP cells, while they remained unaffected in cells from healthy controls. Additionally, lipofermata treatment effectively decreased the expression of *FOXJ1*, a marker of dysregulated ciliogenesis. While the decrease in TP73 expression observed did not reach statistical significance (*P* = 0.057), a trend towards downregulation was evident (Figure 6D). These results collectively indicate that elevated *SLC27A2* expression correlates with increased CRSwNP severity, suggesting its utility as a diagnostic biomarker and its potential as a target for therapeutic intervention in severe manifestations of CRSwNP.

**Figure 6.**
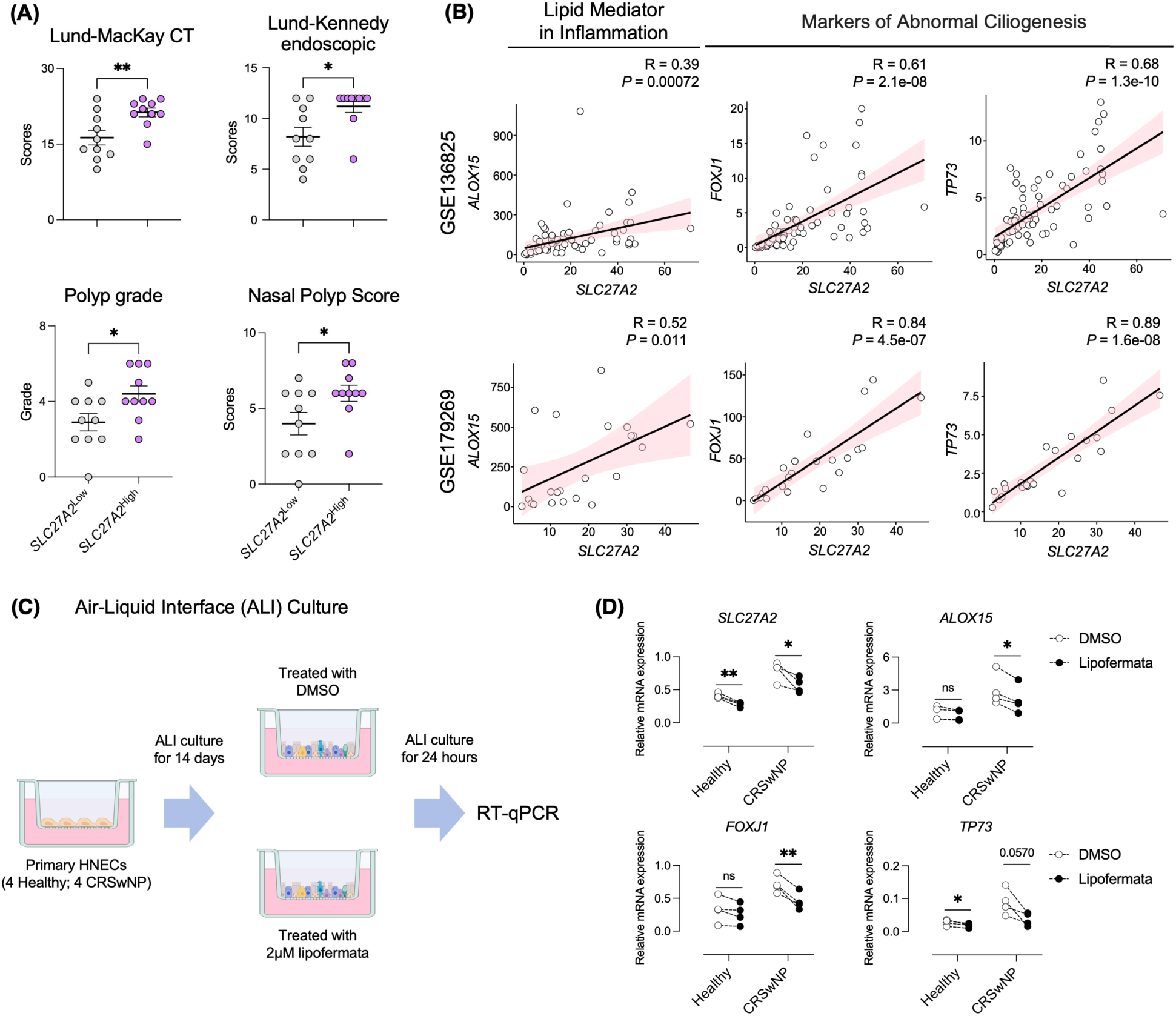
Association of elevated *SLC27A2* expression with severe CRSwNP (A) Comparison of clinical severity indicators (Lund-Mackay CT score, Lund-Kennedy endoscopic score, nasal polyp score, and polyp grade) between *SLC27A2*^Low^ CRSwNP and SLC27A2^High^ CRSwNP groups. (B) Association of *SLC27A2* expression with *ALOX15, FOXJ1*, and *TP73* expression. Datasets GSE136825 (upper panel) and GSE179269 (lower panel) were subjected to Spearman correlation analysis. Correlation coefficients (R) and associated *P*-values are presented. (C) Primary hNECs were cultured using an air–liquid interface (ALI) method and subsequently treated with lipofermata for 24 hours. RNA extraction was performed on these cells, followed by RT-qPCR analysis. (D) mRNA expression of *SLC27A2, ALOX15, FOXJ1*, and *TP73* in ALI-cultured hNECs derived from MT tissue of Healthy (n = 4) and NP tissue of CRSwNP (n = 4) individuals. Cells were treated with or without 2 µM lipofermata for 24 hours, and mRNA levels were assessed via RT-qPCR, normalized to GAPDH mRNA levels. Error bars represent the mean ± SEM. Statistical significance was determined using unpaired or paired t-tests. * P < 0.05, ** P < 0.01, *** P < 0.001, **** P < 0.0001.

## 4. DISCUSSION

CRSwNP is a condition characterized by a complex interplay of inflammatory processes. While it has traditionally been associated with eosinophilia and an increased presence of type 2 inflammatory markers, such as IL-4, IL-5, and IgE, recent insights have broadened our understanding of the pathophysiology of CRSwNP. Notably, the disease exhibits a heterogeneous profile, including not only type 2 but also type 1 (IFN-γ, IL-12) and type 3 (IL-17, IL-22) inflammatory biomarkers. This complexity is further highlighted by the identification of various endotypes, including mixed and unclassifiable forms, which contribute to the diverse clinical manifestations observed in CRSwNP patients.^9,13^ Histological examinations provide additional evidence of this heterogeneity, revealing that up to 40% of CRSwNP patients display a spectrum of infiltrative patterns, encompassing neutrophilic, mixed eosinophilic-neutrophilic, and paucigranulocytic types. These findings underscore the multifaceted nature of CRSwNP and the need for tailored therapeutic approaches.^12,64,65^

In this study, we undertook a comprehensive analysis of transcriptomic profiles within the cellular landscape of nasal polyp tissue. Utilizing integrated bulk and single-cell RNA sequencing, we discerned a pronounced upregulation of genes associated with lipid metabolism in epithelial and myeloid cell populations within the polyp tissue. This finding resonates with the observed higher incidence of CRS in individuals suffering from metabolic syndrome, reinforcing the hypothesis of metabolic disturbances playing a role in nasal polyposis.^22,66–69^ The enhanced synthesis of prostaglandin D2 (PGD2) in CRSwNP and its metabolite delta-12-PGD2, implicated in the recruitment and activation of type 2 T helper cells and eosinophils, exemplifies this metabolic dysfunction.^70^ Elevated levels of unsaturated fatty acids and uric acid in distinct CRSwNP subtypes are indicative of a metabolic environment that fosters neutrophilic inflammation and a type 2 inflammatory response.^69,71–73^ Furthermore, the observed elevation in glucose levels within nasal secretions promotes increased uptake and glycolysis by epithelial cells, accentuating their role in perpetuating inflammation. Our findings articulate the notable overexpression of genes driving lipid peroxidation within the nasal polyp epithelium, which may serve to intensify inflammatory processes and compromise the epithelial barrier, thereby elucidating a mechanistic link between lipid oxidation and CRSwNP pathophysiology.

FATP2, a member of the long-chain fatty acid (LCFA)-coenzyme A ligase family encoded by *SLC27A2*, is significantly overexpressed in the NP epithelium of patients with CRSwNP. Such overexpression is implicated in the dysregulation of lipid and fatty acid metabolism, with FATP2 acting as a critical facilitator for the uptake and integration of polyunsaturated fatty acids (PUFAs) into phospholipids, particularly arachidonic acid (AA) and adrenic acid (AdA).^50^ The absence of FATP2 in cells results in reduced levels of arachidonoyl-phosphatidylethanolamine (AA-PE). ^47^ This highlights the critical role of FATP2 in synthesizing AA-CoA and AdA-CoA, both essential precursors in the lipid peroxidation pathway.^46,47,74^

Positioned at the nexus of PUFA uptake and conversion, FATP2 operates at the apical membrane and endoplasmic reticulum, facilitating the transformation of AA/AdA into their coenzyme A derivatives.^29^ This process is complemented by ACSL4 and LPCAT3 within the ER, promoting the activation and incorporation of AA/AdA-CoA into phosphatidylethanolamine (PE). The subsequent enzymatic action of 15-lipoxygenase, encoded by *ALOX15*, along with the contribution of POR, leads to an accumulation of lipid hydroperoxides, which precipitates an increase in lipid peroxidation and potential cellular injury.^29,56^ Our investigation revealed elevated expression of the enzymes ACSL4, LPCAT3, ALOX15, and POR in *SLC27A2*-positive epithelial cells of NP, compared to *SLC27A2*-negative cells. Moreover, co-localization of FATP2 with markers of lipid peroxidation was established, exhibiting a positive correlation in stained area percentages. In concordance, GO terms related to lipid peroxidation and the mRNA levels of key enzymes, including *ALOX15, DUOX1*, and *POR,* were significantly higher in NP tissue from patients with high *SLC27A2* expression.

While our data suggest a link between FATP2/*SLC27A2* and lipid peroxidation in NP epithelium, the direct contributions of this relationship remain to be fully determined, highlighting an avenue for future research. Notably, the pronounced expression of *SLC27A2* in ciliated cells raises the possibility that the observed loss of these cells in CRSwNP may be attributable to FATP2/*SLC27A2*-mediated lipid peroxidation, presenting an intriguing hypothesis for subsequent exploration.

Our analyses reveal a marked expression of genes linked to ciliopathies in *SLC27A2*-positive cells and in patients with high *SLC27A2* expression in CRSwNP. The aberrant expression of these genes is known to contribute to mucociliary dysfunction, which can facilitate the proliferation of pathogenic bacteria and the formation of biofilms, thereby perpetuating a cycle of chronic inflammation. This aligns with prior findings that associate *FOXJ1* and *TP73* with disrupted ciliary structure and functionality in the epithelium of nasal polyps.^63^ In our cohort, these markers were notably elevated in the group with high *SLC27A2* expression, suggesting a potential mechanistic link between *SLC27A2* overexpression and ciliated cell dysfunction, which may destabilize the epithelial cell environment.

Clinically significant disparities were observed between the *SLC27A2*^High^ and *SLC27A2*^Low^ CRSwNP groups across various disease severity scores, including those derived from Lund-Mackay CT and Lund-Kennedy endoscopic assessments. Therapeutically, the targeted inhibition of *SLC27A2*/FATP2 using lipofermata attenuated the expression of *ALOX15*, implicated in inflammation and lipid peroxidation, and *FOXJ1*, a marker of ciliary disarray, suggesting the modulation of *SLC27A2* as a viable treatment strategy.

In conclusion, our study positions *SLC27A2* as a pivotal biomarker and a potential target for therapeutic intervention in severe CRSwNP. Unbiased transcriptomic profiling has highlighted FATP2/*SLC27A2* as a consistently upregulated entity in CRSwNP, independent of endotypic categorization and intricately associated with lipid peroxidation processes. Inhibition of *SLC27A2* not only resulted in reduced expression of genes central to CRSwNP pathogenesis but also offers a strategic avenue for improving disease prognosis in patients with indeterminate endotypes. These findings advocate for more nuanced patient stratification and personalized treatment approaches, particularly in refractory CRSwNP cases.

## Data Availability

All data produced in the present study are available upon reasonable request to the authors

## Acknowledgements

This work was supported by the National Research Foundation (NRF), Republic of Korea, grant (NRF-2020R1C1C1013939 and NRF-2022R1A2C1011187), which is funded by the Ministry of Science and ICT (MSIT) and grant (2023IL0014) from the Asan Institute for Life Sciences, Asan Medical Center, Seoul, Korea.

