## Supplementary figure legend for "*SLC27A2* as a molecular marker of impaired epithelium in chronic rhinosinusitis with nasal polyps"

**Supplementary Figure 1. Gene expression patterns in nasal polyp tissue.**

(A), Volcano plot demonstrating the transcriptomic expression differences between Healthy and CRSwNP (GSE136825). Genes with a significant FDR < 0.05 and a >two-fold difference in expression level with an average FPKM >2 are represented by colored dots(red, upregulated gene in CRSwNP; blue, downregulated gene in CRSwNP; yellow, lipid metabolic process-related gene).

(B), GO analysis of upregulated genes (red) and downregulated genes (blue) in CRSwNP.

(C), A uniform manifold approximation and projection (UMAP) of the public scRNA-seq dataset (HRA000772) for 72,032 single cells, where points are colored by cell identity.

(D), Dot plot of representative gene expression levels mapped onto each cell type.

(E), Heatmap showing expression levels of lipid metabolic process-related genes that are upregulated in CRSwNP.

**Supplementary Figure 2. Candidate gene expression patterns and lipid accumulation in CRSwNP.**

(A), Feature plots projecting expression patterns of candidate genes.

(B), PCA plots of all known gene expression profiles in public bulk RNA-seq datasets (GSE179269; average FPKM > 1; red, 17 CRSwNP; sky blue, 7 Healthy)

(C), Volcano plot demonstrating the transcriptomic expression differences between Healthy and CRSwNP (GSE179269). Genes with a significant FDR < 0.05 and a >two-fold difference in expression level with an average FPKM >2 are represented by colored dots(red, upregulated gene in CRSwNP; blue, downregulated gene in CRSwNP; yellow, candidate genes).

(D), From the initial set of 6 candidate genes identified using the GSE179269 dataset, a comparison of the expression levels of the 4 genes that were also included in DEGs.

(E), Oil Red O (ORO) staining of Healthy (n = 3) and CRSwNP (n = 7).

Error bar: mean value with ± SEM. P-values were determined by an unpaired *t*-test. * $P < 0.05$, ** $P < 0.01$, *** $P < 0.001$, **** $P < 0.0001$. Scale Bar: 50μm and 100μm.

**Supplementary Figure 3. Comparison of mRNA expression of fatty acid transporter-encoding genes in bulk RNA-seq datasets**

(A), Scatter plot comparing the difference in gene expression levels of fatty acid transporter-encoding genes in CRSwNP and Healthy, drawing on data from GSE179269.

(B), Comparing the expression levels of fatty acid transporter-encoding genes in CRSwNP compared with Healthy, drawing on data from GSE136825 (left) and GSE179269 (right).

(C), Representative IGV genome browser track showing normalized tag density of *ALOX15, IMPA2, and SGPP2*.

Error bar: mean value with ± SEM. P-values were determined by an unpaired *t*-test. * *P* < 0.05, ** *P* < 0.01, *** *P* < 0.001, **** *P* < 0.0001.

**Supplementary Figure 4. Comparative assessment of key features in nECRSwNP and ECRSwNP subtypes**

(A), Comparative Analysis of JESREC scores, blood eosinophil count, tissue eosinophil count, and tissue eosinophil ratio levels in patient samples with nECRSwNP and ECRSwNP. Tissue eosinophil count levels were compared between 6 nECRSwNP and 12 ECRSwNP, while other parameters were compared between 7 nECRSwNP and 13 ECRSwNP.

(B), The mRNA expression of SLC27A2 was measured by RT-qPCR and normalized to GAPDH mRNA in nECRSwNP versus ECRSwNP.

(C), Representative IF staining of DAPI (blue), *SLC27A2*/FATP2 (green), and C11-BODIPY (red) images in nECRSwNP (n = 7) and ECRSwNP (n = 9).

(D and E), Quantification of the percentage area stained by anti-FATP2 (D) and C11-BODIPY (E) was analyzed using the ImageJ.

(F), UMPA plot classified by endotype of CRSwNP. nECRSwNP (left, 41,974 cells) and ECRSwNP (right, 30,058 cells).

(G), Feature plots projecting expression patterns of *SLC27A2*.

(H), Violin plot of the expression levels of *SLC27A2* in nECRSwNP and ECRSwNP.

Error bar: mean value with ± SEM. P-values were determined by an unpaired t-test. * P < 0.05, ** P < 0.01, *** P < 0.001, **** P < 0.0001. Scale Bar: 50μm and 100μm.

**Supplementary Figure 5. Distinct differences between SLC27A2− and SLC27A2+ epithelial cells in NP tissue**

(A), Dot plot of gene expression analyzed by scRNA-seq displaying major markers for the 7 subtypes of epithelial cell.

(B), Pie graph shows the percentage of *SLC27A2*+ cells in NP epithelium

(C), Pie graph shows the percentages of each 7 subtype of epithelial cell in total *SLC27A2*$^{-}$ cells (upper pie graph) and *SLC27A2*$^{+}$ cells (lower pie graph) from CRSwNP patients.

(D), Violin plots showing associated genes with reactive oxygen species metabolic process (left) and positive regulation of lipid oxidation (right).

**Supplementary Figure 6. The transcriptome features of patients with CRSwNP according to the expression level of *SLC27A2*.**

(A), Comparative analysis illustrating the expression levels of representative genes grouped within Cluster 1, drawing on data from GSE179269.

(B), Comparative analysis illustrating the expression levels of representative genes grouped within Cluster 2, drawing on data from GSE136825 and GSE179269.

(C), Comparative analysis illustrating the expression levels of representative genes grouped within Cluster 3, drawing on data from GSE136825 and GSE179269.

Error bar: mean value with ±SEM. P-values were determined by ordinary one-way ANOVA. * $P < 0.05$, ** $P < 0.01$, *** $P < 0.001$, **** $P < 0.0001$.

**Supplementary Figure 7. Association of elevated SLC27A2 expression with endotype and genes related to CRSwNP pathogenesis.0**

(A) Comparative Analysis of JESREC scores, blood eosinophil count, tissue eosinophil count, and tissue eosinophil ratio levels in patient samples with *SLC27A2*$^{Low}$ CRSwNP and *SLC27A2*$^{High}$ CRSwNP. Tissue eosinophil count levels were compared between 9 *SLC27A2*$^{Low}$ CRSwNP and 9 *SLC27A2*$^{High}$ CRSwNP, while other parameters were compared between 10 *SLC27A2*$^{Low}$ CRSwNP and 10 *SLC27A2*$^{High}$ CRSwNP.

(B) Correlations between *SLC27A2* expression levels with *CCL15*, *DUOX1*, *IL5RA*, *NOS2*, and *POR* expression levels. GSE136825 (upper panel) and GSE179269 (lower panel) were analyzed for Spearman correlation. P-values and correlation coefficients (R) were calculated using Spearman correlation analysis.
