## Supplementary figures for "*SLC27A2* as a molecular marker of impaired epithelium in chronic rhinosinusitis with nasal polyps"

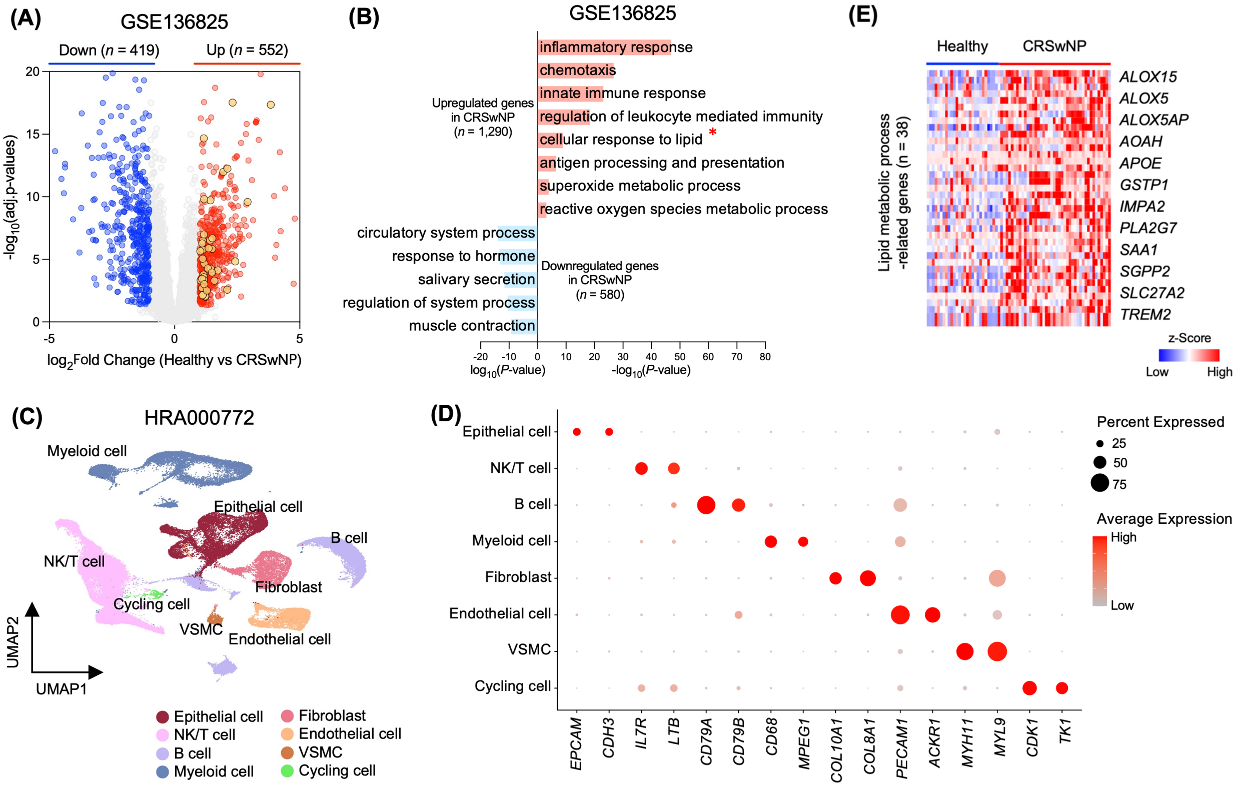


**Supplementary Figure 1_Park et al.**

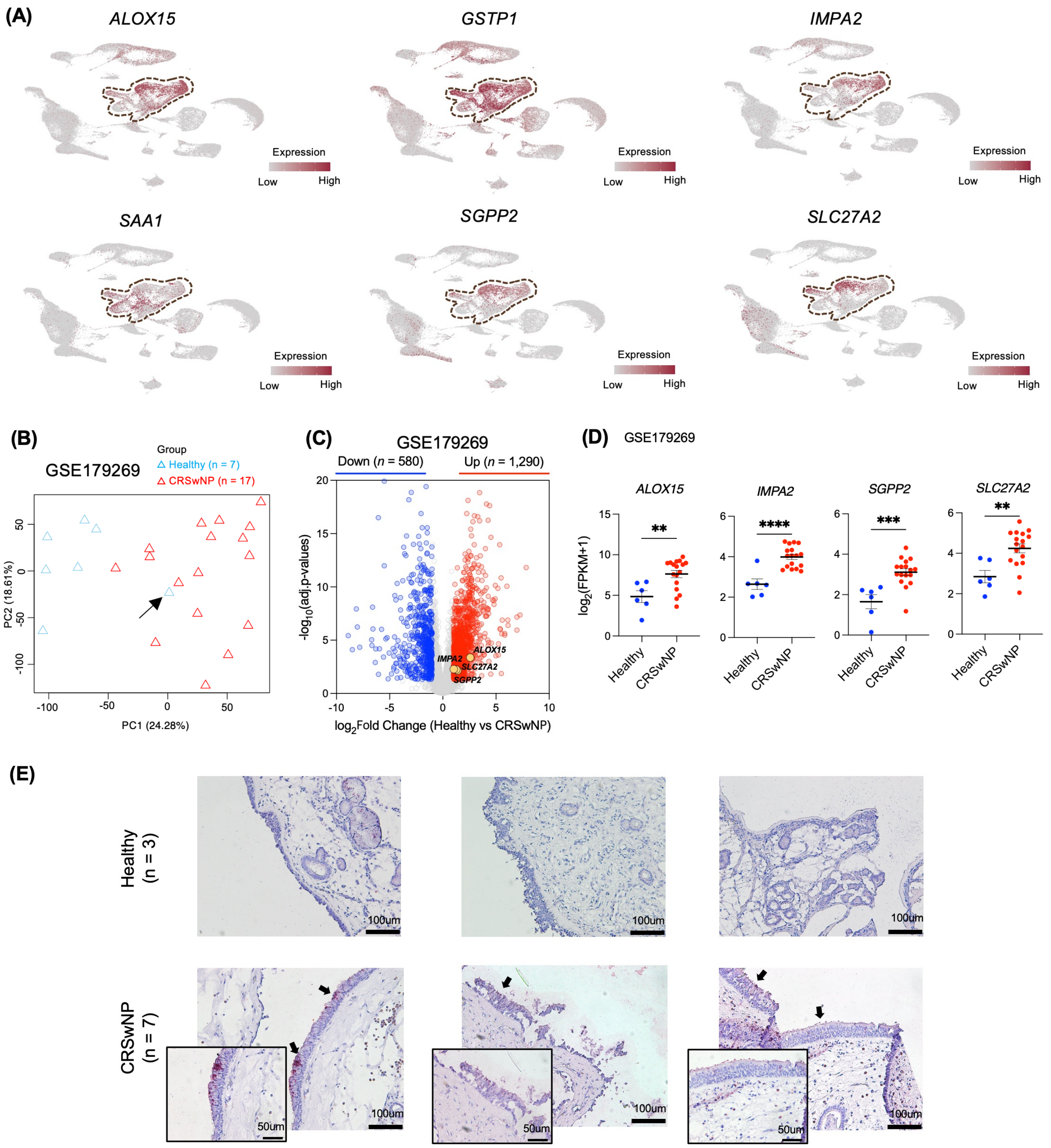


**Supplementary Figure 2_Park et al.**

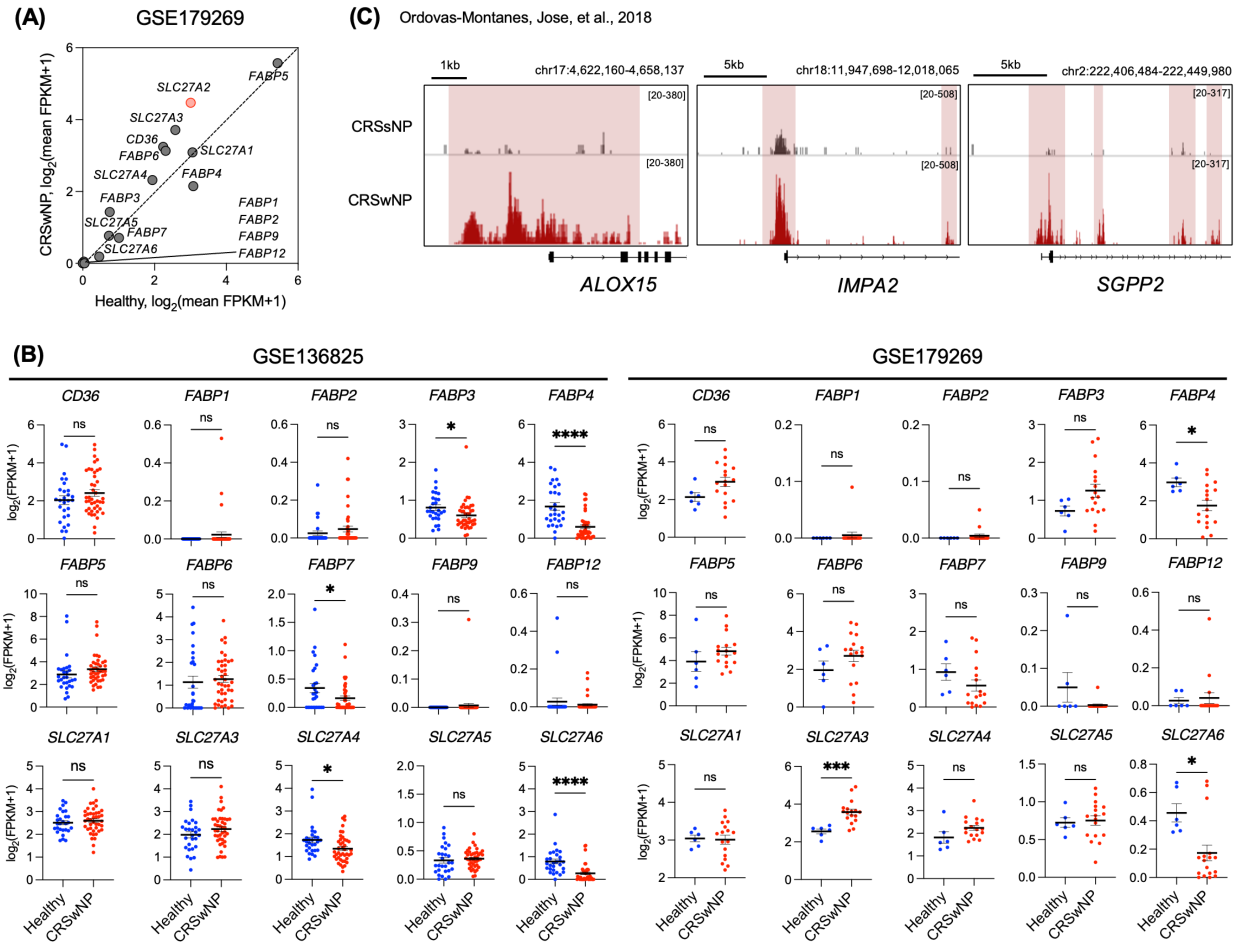


Supplementary Figure 3_Park et al.

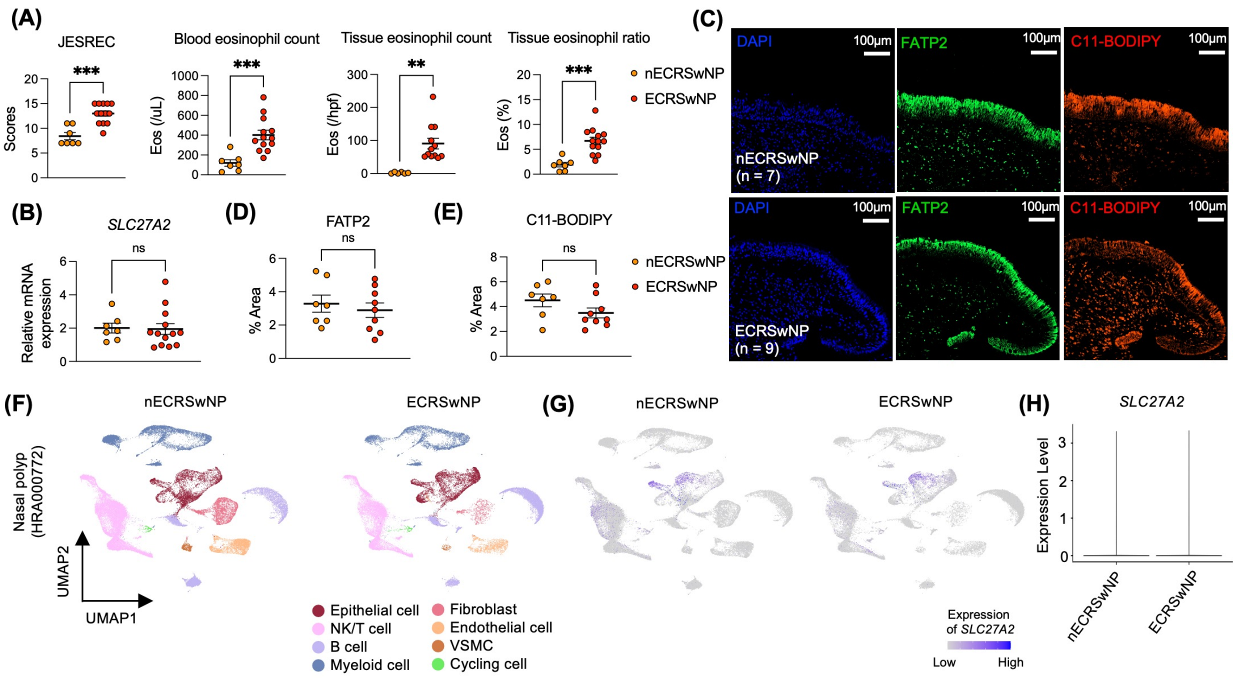


**Supplementary Figure 4_Park et al.**

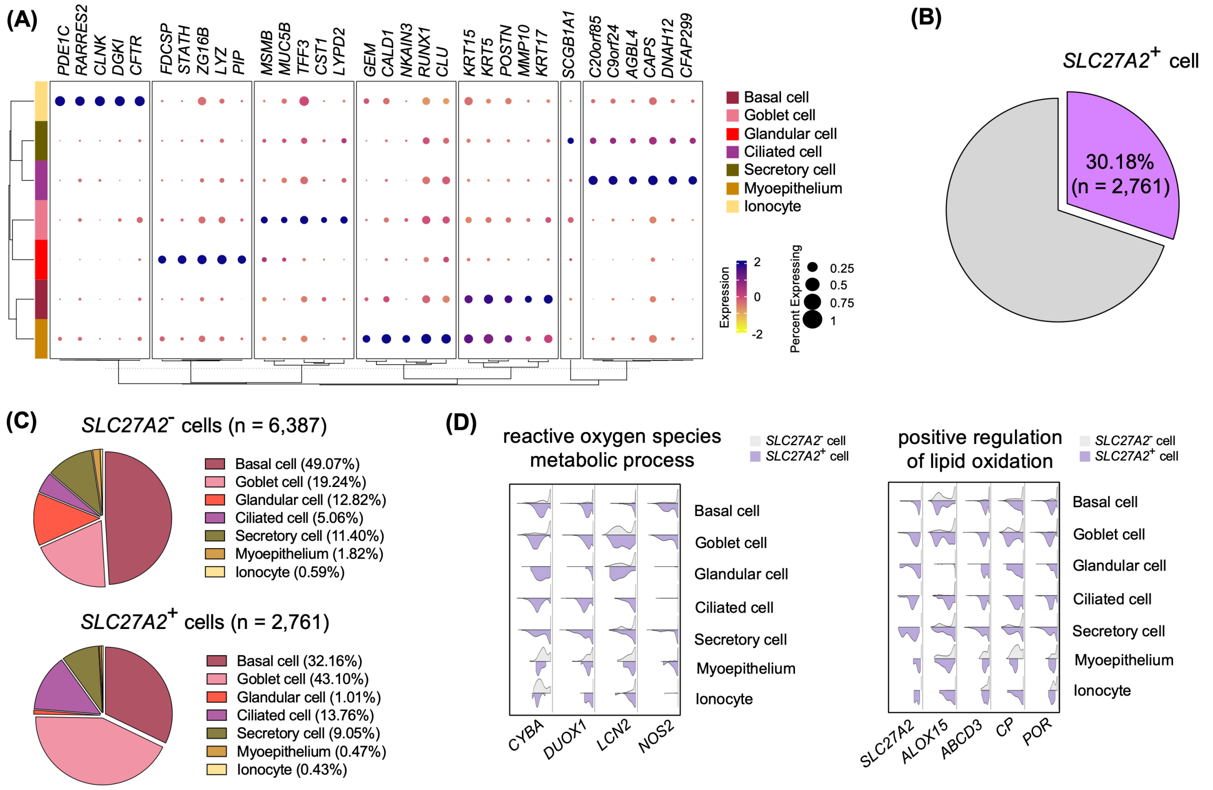


**Supplementary Figure 5_Park et al.**

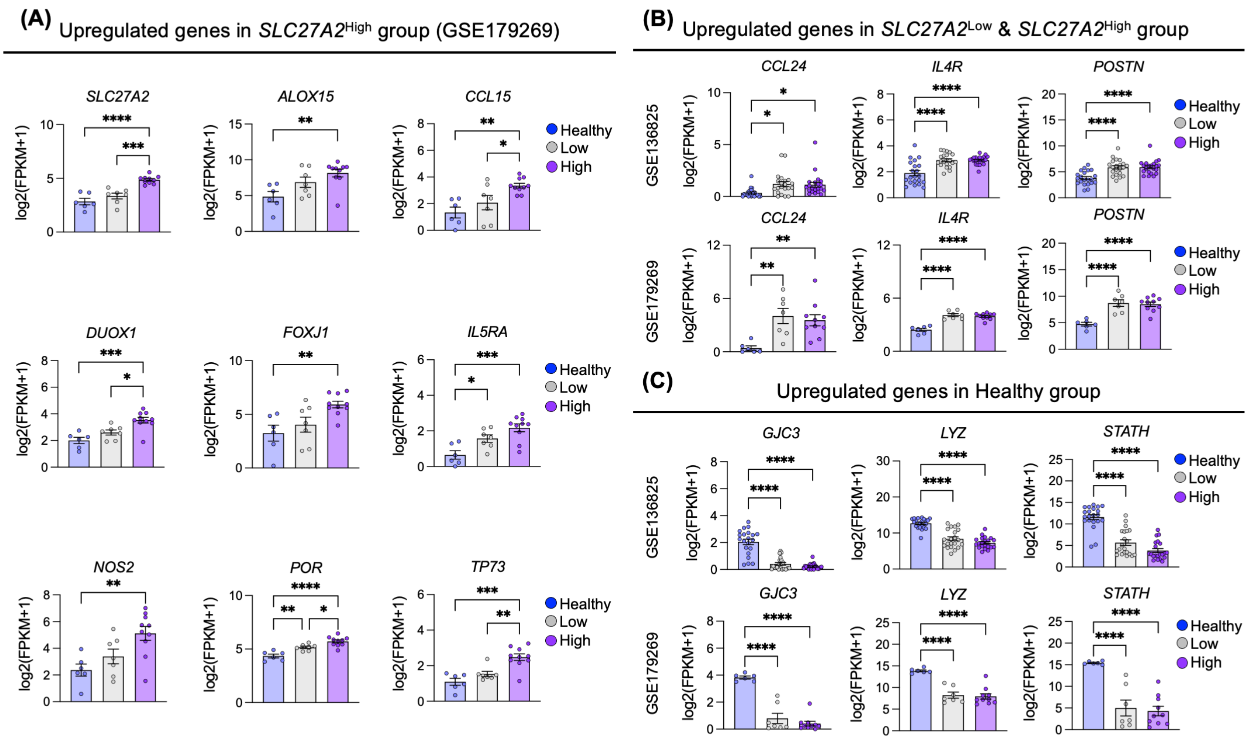


**Supplementary Figure 6_Park et al.**

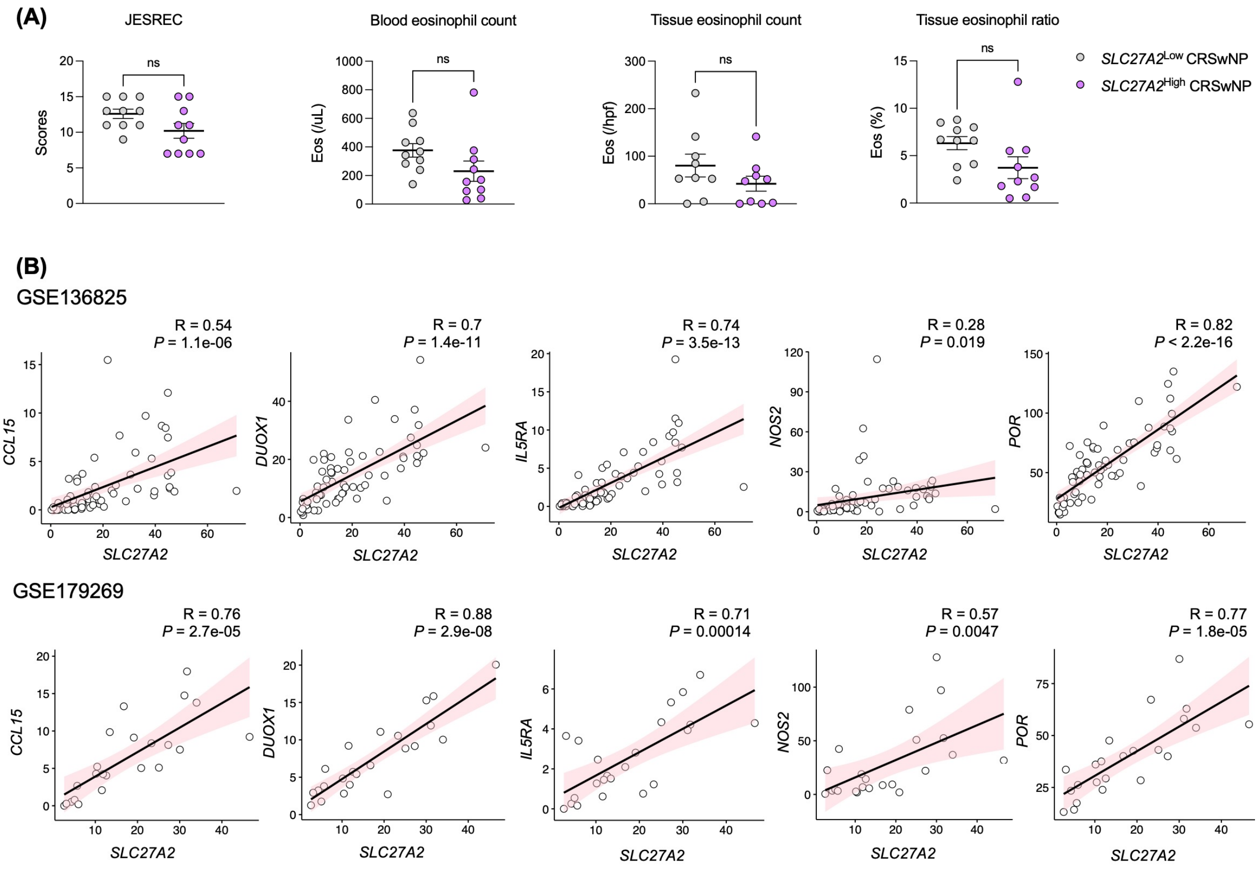


**Supplementary Figure 7_Park et al.**
